# Municipality-level ecological analysis of COVID-19 mortality and infections in Austria, 2020 to 2023

**DOI:** 10.64898/2026.10.15.26365614

**Authors:** Uwe Riedmann, Andrew Stevenson, Chris Dibben, William B. Grant, Martin Sprenger, Lina Zgaga, Johannes Woltsche, Peter Wolf, Winfried März, John PA Ioannidis, Stefan Pilz

## Abstract

**Background:** Previous studies indicated that regional fluctuations in COVID-19 impact were associated with socioeconomic, demographic and environmental factors early in the pandemic. It is unclear whether factors associated with regional COVID-19 mortality and SARS-CoV-2 infections changed over the course of the pandemic.

**Methods:** We evaluated the association of ecological factors with municipality-specific COVID-19 mortality in the national population of Austria between January 2020 and June 2023. Analyses were conducted separately per calendar year and pooled across years via random-effects meta-analysis. Outcomes were analysed using univariable and multivariable zero-inflated negative binomial (ZINB) regression, including annually averaged environmental exposome exposure (including air pollution and UV radiation), socioeconomic, demographic and infection dissemination predictors with significance threshold set to p<0.005. Secondary analysis modelled SARS-CoV-2 infections using of negative-binomial model (NB). Two additional approaches for smaller predictor set selection were applied as sensitivity analyses.

**Results:** Within the entire observation period 18,430 COVID-19 deaths were reported. In univariable analyses (pooled RR (99.5% CI)), COVID-19 mortality was significantly associated with the proportion of commuters (0.79 (0.73-0.86)), residents aged 75-84 (1.18 (1.01-1.37)) and ≥85 years (1.29 (1.04-1.60)), residents with at most compulsory education (1.15 (1.02-1.29)), employed residents aged 15-64 (0.89 (0.85-0.93)), and homeowners (0.91 (0.83-0.99)), as well as with precipitation (0.94 (0.89-0.99)) and the average time with a mean temperature of 5-15 °C (0.95 (0.90-0.99)). In multivariable analyses the share of population aged 85 or older (1.30 (1.09-1.54)) and home ownership (0.92 (0.87-0.98)) were independently significant. Mortality predictor estimates were relatively stable across pandemic years. Across 5,963,882 documented SARS-CoV-2 infections share of employed in 15- to 64-year-olds (1.01 (1.01-1.02)), part-time employment rate (1.01 (1.00-1.02)) and district level nursing care allowance recipiency (0.97 (0.95-0.98)) were associated with detected infections. Overnight stays, population 85 or older, income, compulsory education or less and air pollution all had multiple significant estimates with opposite directionality, leading to non-significant overall associations. Sensitivity analyses outcomes mirrored main analyses outcomes.

**Conclusions:** Throughout the pandemic in Austria, demographic and socioeconomic factors were associated with COVID-19 mortality and infections, but many associations, especially for infections, were unstable over time. These findings underscore the importance of structural vulnerabilities in epidemic impact.

## 1. INTRODUCTION

Associations between socioeconomic, demographic and environmental factors with COVID-19 mortality and SARS-CoV-2 infections have been studied in several countries. Some early studies point towards high age and low socioeconomic status as key factors [1–3] while big seasonal fluctuations in both infections and COVID-19 mortality indicate the potential importance of environmental factors akin to many other diseases [4–11]. However, the relevance of these determinants may have shifted over time with increasing vaccination coverage, repeated infection waves, policy changes and the emergence of new variants.

Ecological evidence on COVID-19 mortality is mixed. Lower socioeconomic status, greater deprivation, higher population density and older age structures are often identified as predictors of COVID-19 mortality [1–3], but associations with temperature [12–14], ultraviolet radiation (UV) [7,12–15], and air pollution [16] vary across settings. This may be due to most studies focusing on early periods and aggregation across years obscuring temporal dynamics, or using very coarse spatial units (e.g countries), where the viability of accounting for international differences is questionable [1–3,17]. Reported infections, while informative about overall exposure, are sensitive to testing availability, policy changes and behavioural changes such as “pandemic fatigue” [18]. It is also questionable how well infection count indicated disease burden given that most severe COVID-19 cases occurred in elderly and highly debilitated people [19]. Prior ecological studies link higher case counts to lower income, younger household composition, and higher density [20], with some studies finding additional environmental associations [21,22], yet few span beyond 2020 in high-testing settings and if they do, they tend to only pool across the entire observation period [17,23]. Given that local policy tightening frequently relied on documented infections [24], distinguishing stable from period-specific determinants of both mortality and infections is essential.

In this ecological analysis of the COVID-19 pandemic in Austria, we retrospectively analysed nationwide data covering the whole pandemic (between January 2020 and June 2023) [25]. We explored the relationship of municipality-specific socioeconomic, demographic and environmental variables using heatmaps and modelled COVID-19 mortality (primary outcome) and SARS-CoV-2 infections (secondary outcome) using zero-inflated negative-binomial (ZINB) and negative-binomial (NB) models for every calendar year, respectively. Both univariable and multivariable analyses were performed and pooled estimates across years were calculated via random-effects meta-analysis.

## 2. METHODS

### 2.1 Study design, procedures, and participants

We conducted a nationwide exploratory ecological study in Austria between January 2020 and June 2023. We used national health data from the Austrian epidemiological reporting system (German: *Epidemiologisches Meldesystem*; EMS) provided by the Austrian Agency for Health and Food Safety (German: *Österreichische Agentur für Gesundheit und Ernährungssicherheit*; AGES), which includes individual data on documented SARS-CoV-2 infections (date), municipality of residence and vital status regarding COVID-19 deaths for every individual with a documented infection [26–28]. Statistics Austria provides a plethora of municipality data to the public (Figure S1) [29]. We extracted municipality data that are aimed to cover important covariates including municipality size (data: total population in 2021), infection dissemination (data: population density in 2023, overnight stays per capita in 2022, percentage of commuters in 2022, work migration balance (ratio of people migrating to the municipality for work versus migrating out of the municipality for work) in 2021 and urbanisation status (urban/semi-urban/rural) in 2025), demographic structure (percentage of 65 to 74 year olds (2022), percentage of 75 to 84 year olds (2022), percentage of 85 year olds or older (2022) and percentage of non-citizens in 2022) and socioeconomic status (data: gross income in 2022, part-time employment rate in 2022, people in training in 2022, employment rate in 15 to 64 year olds in 2022, business units per capita (number of enterprises) in 2022, percentage of home ownership in 2022, percentage of individuals with compulsory education or lower in 2022 and percentage of university graduates in 2022) [29]. Additionally, as a proxy for health status the percentage of individuals with allowance for nursing care (2021) per district was extracted [29]. Environmental data were gathered from multiple different sources. Hourly relative humidity at 2m height and average air temperature (Figure S2 & S3) were available as grid data from the INCA dataset with 1km × 1km resolution [30]. Daily average precipitation rate (Figure S4) measurements were acquired from the Spartacus dataset, a gridded observational dataset for Austria with 1km × 1km resolution [31]. Absolute humidity was calculated from hourly relative humidity and temperature grids using the Magnus formula for saturation vapour pressure [32] (see Supplementary Methods for details; Figure S5 & S6). Monthly measurements of air pollution (i.e., fine particles concentration with diameter ≤ 2.5 μm (PM_2.5_)) were acquired from a dataset that models PM_2.5_ as grid data at a 0.01° × 0.01° resolution using satellite and monitored PM_2.5_ station data (Figure S7) [33]. Grid data of altitude was also publicly available at a 10m × 10m resolution (Figure S8 & S9) [34]. Daily UVA (315-400 nm) and UVB (280-315 nm) measures for Austria were provided by JAXA (Japan Aerospace Exploration Agency) using the MODerate resolution Imaging Spectroradiometer (MODIS) instrument on board NASA’s Aqua and Terra satellites (Figure S10 & S11)[35].

We followed the Strengthening the Reporting of Observational Studies in Epidemiology (STROBE) checklist (Table S1). This is a nationwide study without sample size calculation. Ethical approval was obtained from the ethics committee at the Medical University of Graz, Austria (no. 33-144 ex 20/21). This study was funded by the Austrian Science Fund (FWF) KLI 1188. The analysis was exploratory and shaped by iterative interaction between authors without detailed pre-specified protocol for statistical analysis plan.

### 2.2 Statistical analysis

In this ecological analysis, we calculated Spearman correlations and applied heatmaps to represent correlation structure between different predictors and applied a ZINB model to yearly COVID-19 mortality. Yearly estimates were pooled using a random effects meta-analysis via the “rma” function from the “metafor” package in R. Models that failed to meet pre-specified convergence criteria were refit using a predetermined fallback sequence of alternative model specifications (detailed in the Supplement). Data inspection showed either no (2021 and 2022) or a negligible count (2020 and 2023; 4 each) of municipalities without documented infections. Thus, secondary analyses on infections were performed using the negative-binomial (NB) model only. Analyses were based on municipality level data.

Municipalities were defined as of January 1, 2022 (see supplements for details) [36]. Analyses were performed on a total of 2,115 municipalities, spanning 116 districts (Figure S1). Note that this included city districts of Vienna (on municipality and district level). Throughout the paper we use an alpha of 0.005 as threshold for significance to address the inflated power of a nationwide ecological analysis and multiple comparisons [37]. For Spearman correlations the urbanisation category variable (rural/semi-urban/urban) was treated as an ordered ordinal variable. All continuous predictors were standardised prior to modelling by z-transformation using the R function scale. For mortality outcomes we used 30-day mortality to account for possible mistakes in categorization of deaths. I.e. COVID-19 deaths were only counted if they happened within 30 days of the previously documented SARS-CoV-2 infection [19]. Analyses were performed for individual variables (univariable) and for all variables at once (multivariable).

To account for collinearity we evaluated appropriate values for NB terms using the select method [38]. In this method, predictors that correlate above a threshold (r = 0.7; select07) are inspected and the predictor with the higher variance inflation factor (VIF) is excluded. This is iteratively performed starting with the highest correlation till none of the left-over correlations between predictors is above the set threshold. This approach has shown good variable selection performance in simulation studies [38]. In the select07 based variable selection we used averages across all years for environmental variables and normalized infection count based on population size to asses one set of predictors for all yearly analyses. The subsequent yearly analyses were based on the yearly (non-averaged) values. The ZI term in mortality analysis was based on infection count and income. We used the total population of the respective municipality as offset and included a district-level random effect (Table 1). For a complete list of the variables and used predictors see Table 1. Statistical analyses were performed in R (version 4.5.3) [39].

**Table 1:**
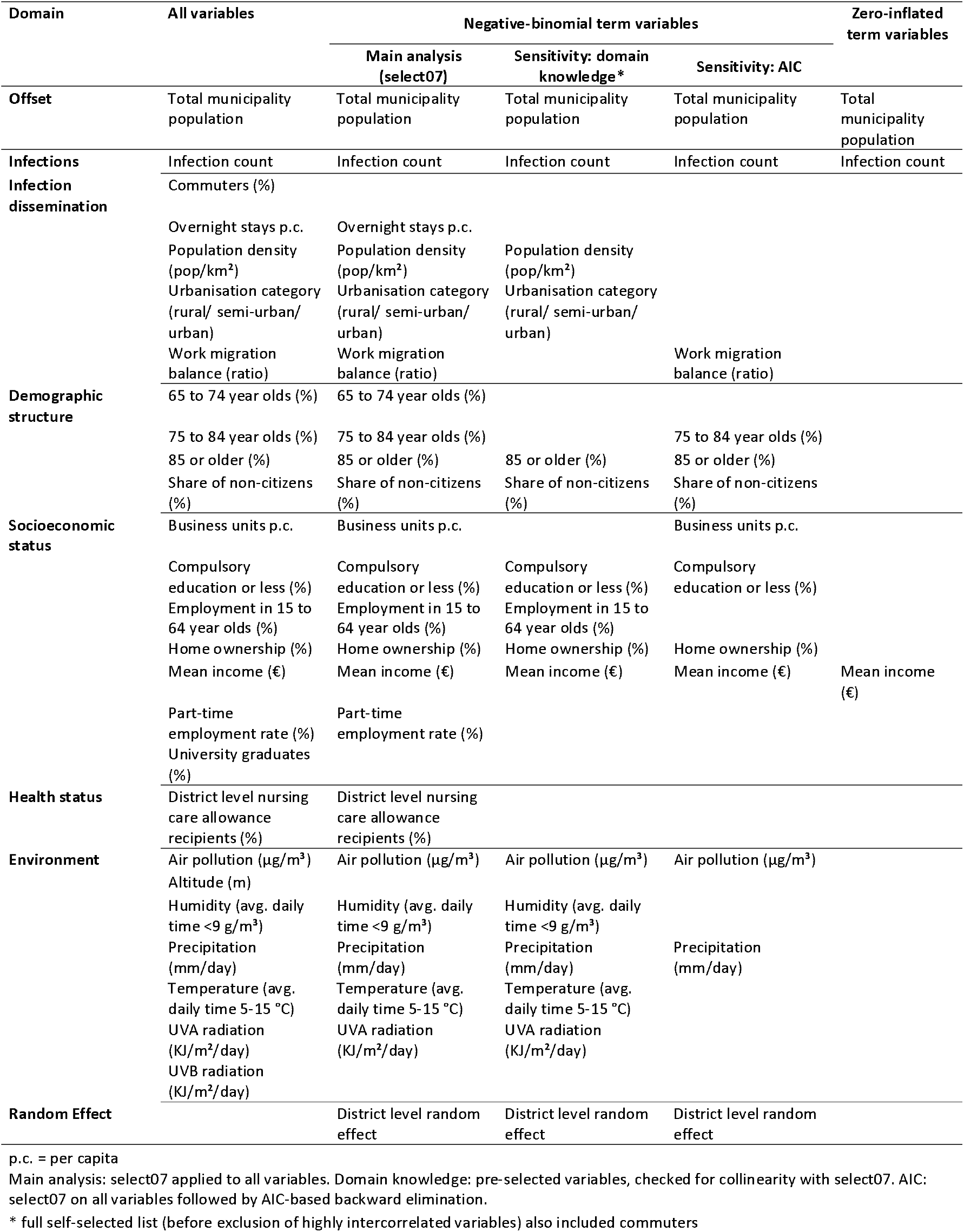
Variables and model predictors by variable-selection approach of primary analysis (COVID-19 deaths).

#### 2.2.1 Sensitivity analyses

We additionally repeated analyses using two more restricted variable-selection approaches. First, we pre-specified a smaller pool of predictors based on domain knowledge of established correlates of SARS-CoV-2 transmission and COVID-19 mortality, and screened this pool for collinearity using the aforementioned select07 approach (Table 1). Second, we applied select07 to the full candidate pool of predictors and then performed AIC-based backward elimination, iteratively removing the predictor whose exclusion most reduced AIC until no further improvement was possible (Table 1). Both approaches are well established for constraining candidate-predictor sets in ecological and environmental epidemiology, where large numbers of correlated predictors relative to available degrees of freedom make unconstrained or fully data-driven selection unstable [40,41]. Both sensitivity models retained the same outcome definitions, exposure offsets, zero-inflation structure, random-effects structure, per-year fitting and meta-analytic pooling as the main analysis. They only differed in the predictor set entered into the model.

### 2.3 Processing

#### 2.3.1 UVA and UVB

Atmospheric absorption and scattering, caused by ozone and water (vapour and cloud) were taken into account by modelling a simplified planetary atmosphere (with a clear atmosphere above a cloud layer). The downward irradiance values, representing the combined direct and diffuse radiation on a horizontal plane for UVA and UVB, were converted to daily figures by incorporating the diurnal cycle of solar zenith angle along with real-time atmospheric conditions. We used Aqua satellite data for all dates except for 17 days for which there were no Aqua satellite data available (September 1 and 2, 2020; April 1 to 15, 2022). For those days we used data from the Terra satellite. If less than 2% of UV satellite raster datapoints were missing on a given day, the missing values were interpolated using the R package “gapfill” [42]. Days with more than 2% missing values were excluded from the analyses (see supplements for details). UVA and UVB data had systematic missing data on the same dates. On this basis we included/excluded 356/10 days, 342/23 days, 298/66 and 179/2 for 2020, 2021, 2022 and 2023, respectively. We estimated mean daily UVA and UVB values of municipalities for every interval individually.

#### 2.3.2 Other environmental variables

For precipitation and pollution we estimated averaged values (per municipality and time period; Figures S4 & S7). Temperature and absolute humidity were considered based on ranges previously found to be associated with higher COVID-19 disease spread. Specifically we used municipalities average hours per day between 5° C and 15° C for temperature (Figure S3) and average hours per day below 9 g/m³ for absolute humidity (Figure S6) [43]. For municipality values of altitude, we used the 10^th^ percentile altitude value of the respective municipality to address possible skewed mean values in mountainous regions (Figure S8 and S9).

#### 2.3.3 Socioeconomic variables

Some socioeconomic factors were only available for the most recent municipality stratification (two municipalities merged in 2025). In these cases, we used values from the now merged municipality for both previous municipalities (e.g. urbanisation category). See more details in the supplements.

## 3. RESULTS

Between January 2020 and July 2023, a total of 18,430 COVID-19 deaths (30-day mortality) and 5,963,882 SARS-CoV-2 infections with plausible municipality data were documented. Due to missing or wrong municipality data 38,156 infections and 185 deaths were excluded. In 2020, 2021, 2022 and 2023, we recorded 7,058, 6,638, 4,171 and 563 COVID-19 deaths and 354,557, 904,660, 4,336,472 and 368,193 SARS-CoV-2 infections, respectively.

### 3.1 Heatmaps

Most variables correlated significantly with most other variables showing the complex relationship between variables (Figure 1). Most infection dissemination and socioeconomic variables were associated with population size. Age structure variables were mostly related to population density, employment rate in 15 to 64 year olds and nursing care allowance recipiency as well as number of infected per capita. Most environmental variables were relatively stable across years and were highly correlated to each other (Figure S13). Notably, 2023 only covered half the year, which did not noticeably affect relative exposure in any environmental variables except for air pollution (Figure S13).

**Figure 1:**
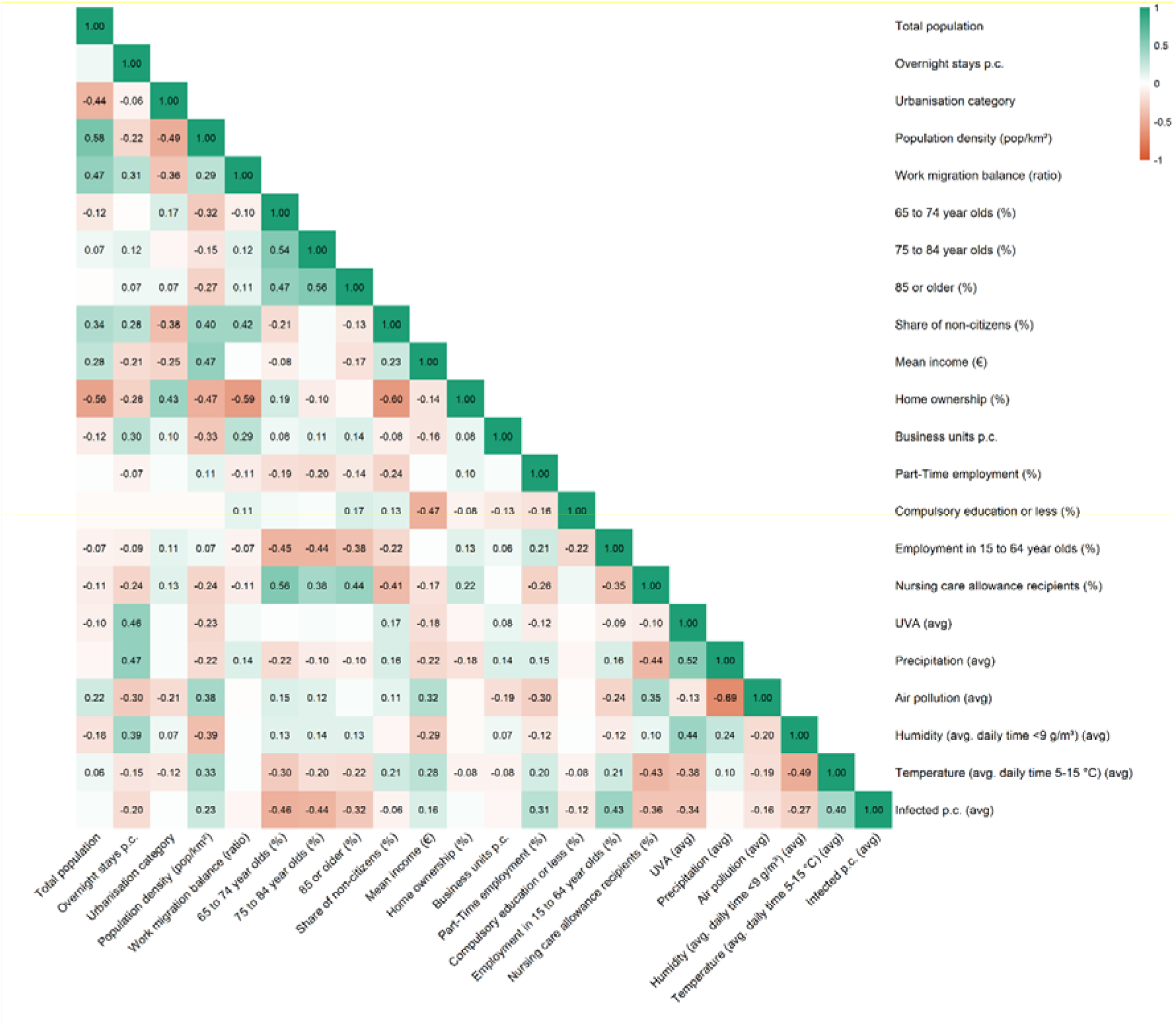
Heatmap of all considered variables, with 2022 values of environmental variables. Only significant (p < 0.005) Spearman correlations are shown.

#### 3.1.1 Selected variables

The select07 process dropped the variables altitude (due to correlation with pollution, precipitation and UVB), UVB (due to correlation with UVA), percentage of commuters (due to correlation with employment rate in 15 to 64 year olds) and share of university degrees (due to correlation with income). Non-dropped variables should be interpreted as a composite version of the highly correlated variables that were dropped [38].

### 3.2 COVID-19 mortality

#### 3.2.1 Univariable analyses

Pooled univariable estimates of COVID-19 mortality are positively associated with 75 to 84 year olds, 85 or older and compulsory education or less, as well as negatively associated with commuters, employment in 15 to 64 year olds, home ownership, precipitation and temperature (avg. daily time 5-15 °C) (Table 2). Multiple predictors had at least one significant yearly estimate (Table S2). Between-year heterogeneity was moderate to high in most predictors (I² > 50%) (Table S3).

**Table 2:** Pooled univariable associations of each predictor with COVID-19 deaths and SARS-CoV-2 infections.

| Domain | Variable | Deaths (primary analysis) |  | Infections (secondary analysis) |  |
| --- | --- | --- | --- | --- | --- |
|  |  | RR (99.5% CI) | p | RR (99.5% CI) | p |
| **Infections** | Infection count | 1.02 (0.99–1.05) | 0.11 | – | – |
| **Infection dissemination** | Commuters (%) | **0.79 (0.73–0.86)** | **<0.001** | **1.04 (1.01–1.08)** | **0.001** |
|  | Overnight stays p.c. | 0.97 (0.91–1.03) | 0.16 | 0.99 (0.97–1.02) | 0.42 |
|  | Population density (pop/km<sup>2</sup>) | 1.00 (0.97–1.03) | 0.95 | 1.01 (0.95–1.08) | 0.54 |
|  | Urbanisation category (rural/ semi-urban/ urban) | 1.13 (0.91–1.42) | 0.11 | 1.01 (0.93–1.11) | 0.67 |
|  | Work migration balance (ratio) | 1.06 (0.97–1.15) | 0.055 | 1.01 (0.97–1.06) | 0.47 |
| **Demographic structure** | 65 to 74 year olds (%) | 1.09 (0.94–1.27) | 0.11 | 0.97 (0.93–1.00) | 0.007 |
|  | 75 to 84 year olds (%) | **1.18 (1.01–1.37)** | **0.002** | 0.98 (0.92–1.04) | 0.31 |
|  | 85 or older (%) | **1.29 (1.04–1.60)** | **0.001** | 1.00 (0.95–1.05) | 0.84 |
|  | Share of non-citizens (%) | 1.06 (0.96–1.17) | 0.096 | 0.99 (0.94–1.06) | 0.81 |
| **Socioeconomic status** | Business units p.c. | 1.00 (0.97–1.03) | 0.98 | 1.00 (0.99–1.01) | 0.64 |
|  | Compulsory education or less (%) | **1.15 (1.02–1.29)** | **0.001** | 1.00 (0.94–1.06) | 0.92 |
|  | Employment in 15 to 64 year olds (%) | **0.89 (0.85–0.93)** | **<0.001** | 1.02 (0.98–1.06) | 0.10 |
|  | Home ownership (%) | **0.91 (0.83–0.99)** | **0.001** | 0.99 (0.92–1.07) | 0.68 |
|  | Mean income (€) | 0.93 (0.86–1.01) | 0.017 | 1.01 (0.89–1.15) | 0.81 |
|  | Part-time employment rate (%) | 0.96 (0.91–1.00) | 0.007 | 1.01 (0.98–1.04) | 0.28 |
|  | University graduates (%) | 0.98 (0.91–1.05) | 0.31 | 1.01 (0.89–1.15) | 0.75 |
| **Health status** | District level nursing care allowance recipients (%) | 1.12 (0.90–1.38) | 0.15 | **0.95 (0.92–0.97)** | **<0.001** |
| **Environment** | Air pollution (µg/m<sup>3</sup>) | 1.04 (0.97–1.13) | 0.10 | 1.00 (0.90–1.12) | 0.95 |
|  | Altitude (m) | 0.97 (0.90–1.04) | 0.24 | 0.96 (0.83–1.12) | 0.48 |
|  | Humidity (avg. daily time <9 g/m<sup>3</sup>) | 0.99 (0.95–1.04) | 0.74 | 0.99 (0.97–1.02) | 0.60 |
|  | Precipitation (mm/day) | **0.94 (0.89–0.99)** | **0.002** | 0.98 (0.89–1.08) | 0.60 |
|  | Temperature (avg. daily time 5-15 °C) | **0.95 (0.90–0.99)** | **0.002** | 1.03 (0.93–1.14) | 0.37 |
|  | UVA radiation (KJ/m<sup>2</sup>/day) | 0.95 (0.83–1.08) | 0.28 | 0.98 (0.92–1.05) | 0.51 |
|  | UVB radiation (KJ/m<sup>2</sup>/day) | 0.95 (0.84–1.09) | 0.31 | 0.97 (0.88–1.08) | 0.46 |

RR = rate ratio per 1 SD increase in the (standardised) predictor; for the categorical urbanisation variable, the first non-reference level versus the reference level. 99.5% CI.
Bold: p < 0.005. p. c. = per capita.

#### 3.2.2 Multivariable analyses

Meta analysis of COVID-19 mortality models showed significant associations with the share of population aged 85 or older (pooled RR: 1.30 (99.5% CI: 1.09–1.54)) and inversely associated with home ownership (pooled RR: 0.92 (99.5% CI: 0.87–0.98)) (Table S4 & Figure 2). In both cases outcome directionality did not differ between years, but not all yearly associations were significant (Table S5 & Figure 2). Share of population aged 85 or older and multiple other predictors had high between-year heterogeneity (Table S5). Yearly analyses also showed significant associations with mean income, percentage of individuals with compulsory education or lower and precipitation for a single year each, but no overall statistically significant effect in the meta-analysis (Table S5 & Figure 2). Explained variance of multivariable models (R²) can be seen in Table S6 and S7. Note that R² values are not directly comparable between deaths and infections, because rare events such as deaths are subject to much more random variation than the very frequent infections, which lowers R² regardless of how well the model fits.

**Figure 2:**
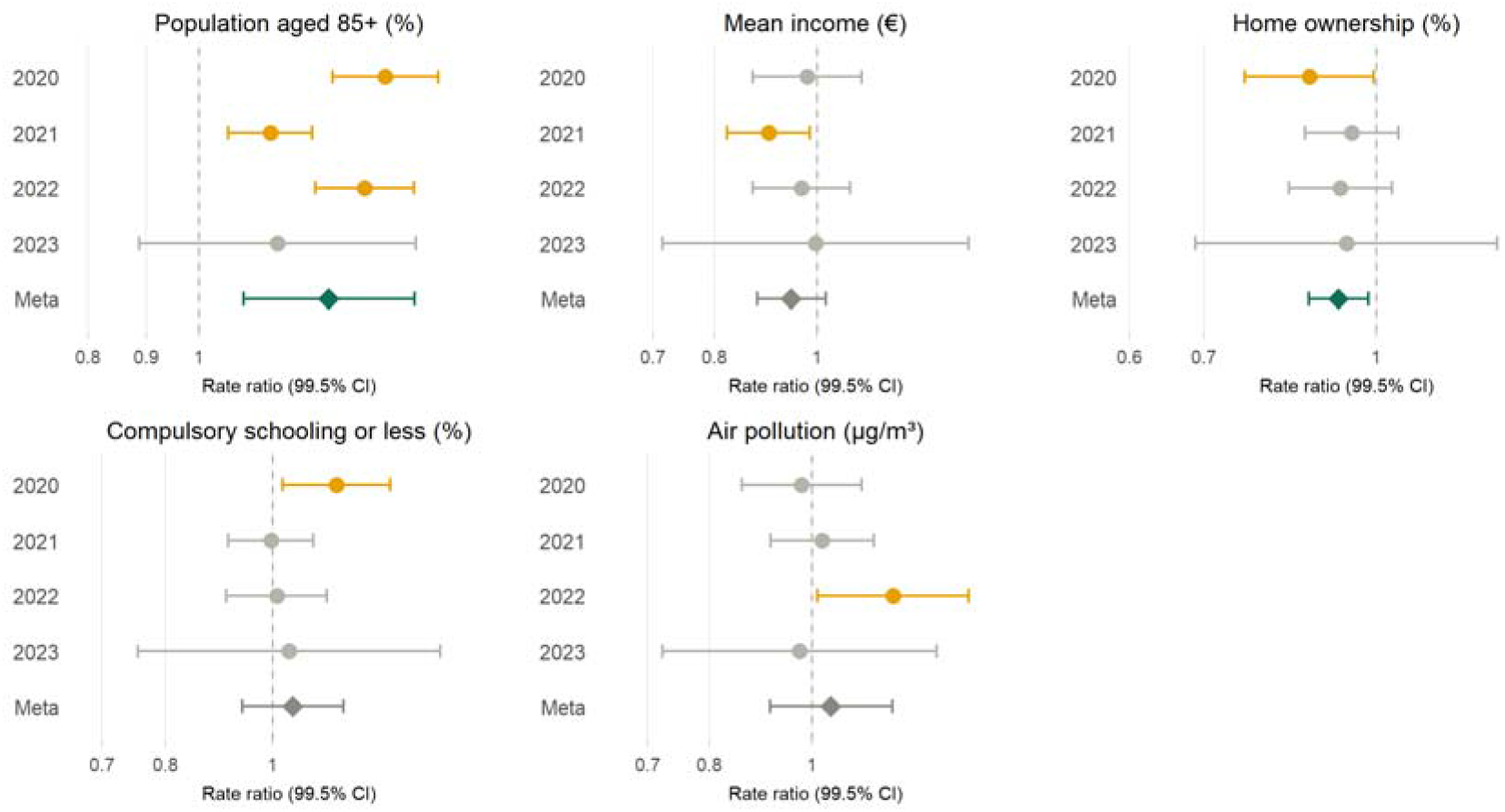
Multivariable yearly and meta-analysis outcomes for adjusted RRs of predictors of COVID-19 mortality with at least one significant estimate (p < 0.005). See Table S4 and S5 for all results.

### 3.3 SARS-CoV-2 infections

#### 3.3.1 Univariable analyses

Pooled univariable estimates of SARS-CoV-2 infections were positively associated with commuters and negatively associated with district level nursing care allowance recipients (Table 2). Most predictors had at least one significant yearly estimate (Table S8) and between-year heterogeneity was very high across the board, in all predictors but the percentage of district level nursing care allowance recipients and business units per capita (Table S3).

#### 3.3.2 Multivariable analyses

Meta analysis of SARS-CoV-2 infection models showed significant associations with share of employed in 15- to 64-year-olds (pooled RR: 1.01 (99.5% CI: 1.01–1.02)), part-time employment rate (pooled RR: 1.01 (99.5% CI: 1.00–1.02)) and district level nursing care allowance recipiency (pooled RR: 0.97 (99.5% CI: 0.95–0.98)) (Figure 3 & Table S9). Multiple other variables showed significant associations with infection count for specific years (Figure 3 & Table S10), with on average 7.5 significant predictors per yearly analysis. Overnight stays, population 85 or older, income, compulsory education or less and air pollution all had multiple significant estimates with opposite directionality (Figure 3). Overall, most predictors had very high between-year heterogeneity (I² > 75%) (Table S9).

**Figure 3:**
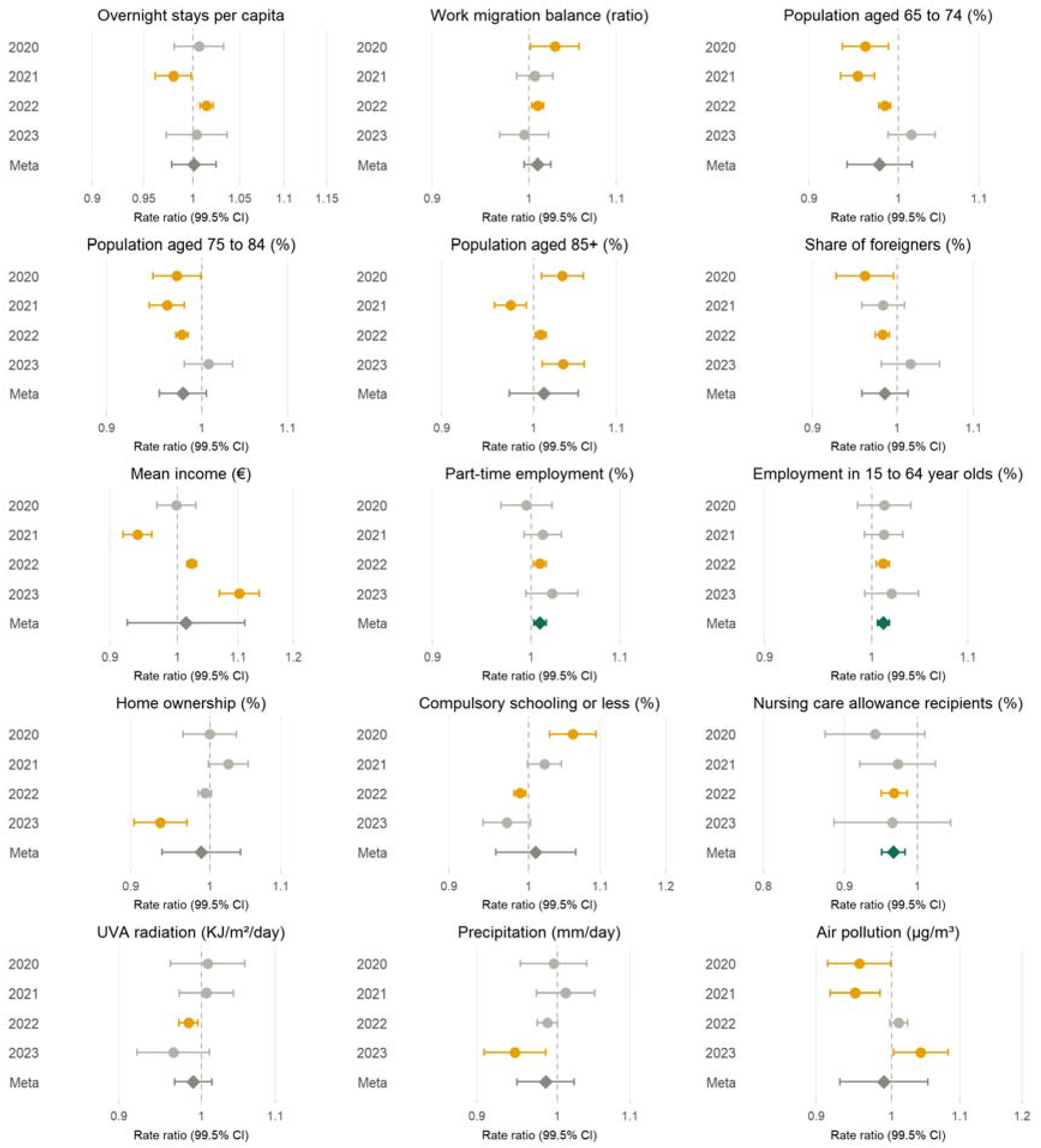
Multivariable yearly and meta-analysis outcomes for predictors of SARS-CoV-2 infections with at least one significant estimate (p < 0.005). See Table S6 and S7 for all results.

### 3.4 Sensitivity Analyses

In alternative predictor selection of multivariable analysis, both self-selected and AIC based predictor selections found significant associations for COVID-19 mortality with the share of population aged 85 or older and inverse association with home ownership (Table S4, Figure S14). Self-selected additionally found a significant inverse association with the share of non-citizen (Table S4). SARS-CoV-2 infections only showed consistent associations across different selections for employment in 15 to 64 year olds (Tables S9). Self-selected additionally found a significant inverse association with share of non-citizens. The associations with part-time employment rate and district level nursing care allowance recipients found in the main analysis could not be replicated in the sensitivity variable selections.

## 4. DISCUSSION

We conducted a nationwide, municipality-level, time-stratified ecological analysis of Austria (2020–2023). Across years, multivariable analyses showed that COVID-19 mortality was associated with older age structure (share aged 85+) and lower percentage of home ownership, with the direction of estimates largely stable. While pooled analysis documented SARS-CoV-2 infections was significantly association with higher share of employed in 15- to 64-year-olds, higher part-time employment rate and lower district level nursing care allowance, yearly analyses showed many additional predictors that starkly varied in directionality by year. Several covariates (share of 85+ year olds population age, mean income, pollution and compulsory schooling) changed direction between early (2020–2021) and later (2022–2023) pandemic periods. Annually averaged environmental factors did not show consistent associations with COVID-19 mortality or infections, and infection count did not show any association with COVID-19 mortality in our model. Between-year heterogeneity was high, suggesting that period-specific context substantially affected observable associations.

In multivariable analyses, the positive association of mortality with older age structure and the inverse association with home ownership (as an indicator of socioeconomic status) align with prior ecological evidence on COVID-19 severity and deprivation [3]. Stability of these relationships despite changing variants, vaccination rollout, and clinical improvements suggests that structural vulnerability at the municipal level remained salient determinants of COVID-19 mortality risk. However, note that home ownership was only significant in the yearly analysis of 2020 and pooled estimates. Similarly, mean income was only significant in 2021 but also consistently points towards a negative association of socioeconomic status and COVID-19 mortality. The lack of an independent association of mortality with documented infections might reflect decoupling of case detection from severe outcomes due to differential testing practices, testing performed mostly in non-risk populations and vaccine-mediated disease attenuation, as well as lag and potential measurement issues [24].

The pooled analysis of predictors suggests that the most robust correlate of higher documented infection rates was likely testing conducted predominantly in non risk populations (higher share of employed in 15- to 64-year-olds and higher part-time employment rate). Austria had an exceptionally high testing volume and long periods of free available testing combined test certification requirements for social events. During the pandemic many workplaces may have opt to frequently test their workforce leading a relatively higher documented infection count in regions with higher employment. Most other predictors were highly time-dependent. The substantial between-year heterogeneity and direction reversals for variables like income and pollution point to shifting testing and exposure mechanisms during the pandemic. Variation in testing policies, incentives (e.g. certification requirements), pandemic fatigue and increase in tourism towards the end of the pandemic have likely contributed to the heterogeneity of yearly estimates. Also, while many elderly people may have been more protected in the early years, eventually almost all elderly people did get infected, but in later pandemic phases with low risk of disease complications. Importantly, all outcomes in analysis of documented infections were small. Overall, these findings indicate that reported cases may have been a suboptimal indicator of population risk at the ecological scale.

Despite prior literature linking temperature, UV, and air pollution to COVID-19 metrics [7,9,15,23,44], we found univariate protective effects for precipitation and temperature (avg. daily time 5-15 °C) only, and no robust environmental associations in multivariable analyses. This could be due to multiple factors. Environmental analyses are restricted by the high correlation of the factors. As our approach analyses annual environmental exposures, effects that differ depending on time of year and infection waves cannot be identified. E.g., UV exposure may be most relevant in darker months, when underexposure is most certain [7]. Austria also had rather small variations in average regional environmental fluctuations mainly coinciding with differences in altitude which may further coincide with potentially relevant factors not accounted for such as health care accessibility.

Sensitivity analyses further strengthen our findings by indicating that independent of variable selection approach, multivariable analyses find age structure and socioeconomic status proxies to be the only relevant indicators in COVID-19 deaths (Table S4). None of the variable selections found a significant association with SARS-CoV-2 infections. Secondary analyses further underline the instability of predictors of SARS-CoV-2 infections as employment in 15 to 64 year olds was the only significant predictor across all analyses (Table S9). Infections may have been too dependent on policy changes, testing rates and other unmeasured local drives to be strongly and consistently associated with any fixed factor.

The study is limited by its ecological nature and the quality of the available data. Ecological conclusions cannot be directly attributed to individuals [45]. While we account for multiple demographic and socioeconomic variables, there is the possibility that we missed crucial factors in our analysis. For example, we do not have data on healthcare access or nursing home capacities per municipality. Influencing factors also include discrepancies between documented and actual infections as well as under- and overcounting of COVID-19 death counts [46–49]. For socioeconomic and demographic variables, we use the same values for all yearly analyses, and if no other values are available, even values that are beyond the observation period (e.g. urbanisation categorization from 2025). We believe this is a minor issue as changes in these categories are expected to be minor over the time period of our study and thus likewise negligible for our findings. Another limitation may be the fact that 2023 measurements only cover half the year (as testing stopped after June 2023). We also didn’t investigate specific derivable mediators between ambient environmental factors and COVID-19 outcomes such as Vitamin D [50]. It has to be noted that ecological designs with annual averages restrict consistently changing environmental conditions to a single yearly value. Thus, this study design is limited in its power to identifying the impact of temporally changing environmental factors. Lastly, while we set the alpha value to 0.005 to attenuate the inflated power of nationwide ecological studies and multiple comparisons, many significant estimators (especially in the infection prediction model) are potentially still statistically significant due to the high outcome count but may be irrelevant for public health practice.

Across four pandemic years, mortality was consistently associated with older age structure and lower home ownership, whereas reported infections were associated with share of employed in 15- to 64-year-olds, higher part-time employment rate and lower share of nursing care allowance recipients. Yearly analyses showed high period-specific fluctuations in relevant predictors of documented SARS-CoV-2 infections. Environmental variables showed no robust independent associations in this Austrian ecological context, possibly due to limited seasonality dependent environmental effects not assessed in the present study. These results support caution in relying on case counts for local decision-making and highlight the importance of structural vulnerability and more robust outcome indicators when monitoring epidemic impact.

## Data Availability

The data that support the findings of this study are available upon request with approval needed from the Austrian Agency for Health and Food Safety (AGES), Vienna, Austria and Statistics Austria respectively. The data are not publicly available due to restrictions pertaining to contained information that could compromise the privacy of patients.

## Contributors

UR and SP conceptualized the study with contributions by JPAI. UR wrote the original draft. UR performed the formal analyses and visualization with contributions of AS. UR and AS were involved in data curation. SP was supervising. All authors contributed to writing, reviewing, and editing the manuscript, and approved the final version before submission.

## Funding

This study was funded by the Austrian Science Fund (FWF) KLI 1188.

## Declaration of interests

The authors declare no conflict of interests.

## Acknowledgements

The authors thank all persons and organizations involved in data collection, and to those who supported the identification and acquisition of relevant datasets.

## Notes

### Competing Interest Statement

The authors have declared no competing interest.

### Author Declarations

Ethical approval was obtained from the ethics committee at the Medical University of Graz, Austria (no. 33-144 ex 20/21)

