## Supplements for "Municipality-level ecological analysis of COVID-19 mortality and infections in Austria, 2020 to 2023"

#### *Table S1:* STROBE Statement—Checklist of items for *cohort studies*

|  | **Item No** | **Recommendation** | **Main text page** |
| --- | --- | --- | --- |
| **Title and abstract** | 1 | (*a*) Indicate the study’s design with a commonly used term in the title or the abstract | Title, page 1 |
|  |  | (*b*) Provide in the abstract an informative and balanced summary of what was done and what was found | Page 3 & 4 |
| **Introduction** | | |  |
| Background/rationale | 2 | Explain the scientific background and rationale for the investigation being reported | Page 5 & 6 |
| Objectives | 3 | State specific objectives, including any prespecified hypotheses | Page 6 |
| **Methods** | | |  |
| Study design | 4 | Present key elements of study design early in the paper | Page 6 to 8 |
| Setting | 5 | Describe the setting, locations, and relevant dates, including periods of recruitment, exposure, follow-up, and data collection | Page 6 to 11 |
| Participants | 6 | (*a*) Give the eligibility criteria, and the sources and methods of selection of participants. Describe methods of follow-up | Page 6 to 11 |
|  |  | (*b*) For matched studies, give matching criteria and number of exposed and unexposed | n/a |
| Variables | 7 | Clearly define all outcomes, exposures, predictors, potential confounders, and effect modifiers. Give diagnostic criteria, if applicable | Page 6 to 11 |
| Data sources/ measurement | 8* | For each variable of interest, give sources of data and details of methods of assessment (measurement). Describe comparability of assessment methods if there is more than one group | Page 5 to 7 |
| Bias | 9 | Describe any efforts to address potential sources of bias | Page 8 to 10 |
| Study size | 10 | Explain how the study size was arrived at | Page 7 |
| Quantitative variables | 11 | Explain how quantitative variables were handled in the analyses. If applicable, describe which groupings were chosen and why | Page 6 to 11 |
| Statistical methods | 12 | (*a*) Describe all statistical methods, including those used to control for confounding | Page 8 to 10 |
|  |  | (*b*) Describe any methods used to examine subgroups and interactions | Page 8 to 10 |
|  |  | (*c*) Explain how missing data were addressed | Page 6 to 11 & Supplements |
|  |  | (*d*) If applicable, explain how loss to follow-up was addressed | n/a |
|  |  | (*e*) Describe any sensitivity analyses | Page 9 & 10 |
| **Results** | | |  |
| Participants | 13* | (a) Report numbers of individuals at each stage of study—eg numbers potentially eligible, examined for eligibility, confirmed eligible, included in the study, completing follow-up, and analysed | Page 11 |
|  |  | (b) Give reasons for non-participation at each stage | n/a |
|  |  | (c) Consider use of a flow diagram | n/a |
| Descriptive data | 14* | (a) Give characteristics of study participants (eg demographic, clinical, social) and information on exposures and potential confounders | Page 10 & 11, & Supplements |
|  |  | (b) Indicate number of participants with missing data for each variable of interest | n/a |
|  |  | (c) Summarise follow-up time (eg, average and total amount) | n/a |
| Outcome data | 15* | Report numbers of outcome events or summary measures over time | Page 11 |
| Main results | 16 | (*a*) Give unadjusted estimates and, if applicable, confounder-adjusted estimates and their precision (eg, 95% confidence interval). Make clear which confounders were adjusted for and why they were included | Page 11 to 14 |
|  |  | (*b*) Report category boundaries when continuous variables were categorized | n/a |
|  |  | (*c*) If relevant, consider translating estimates of relative risk into absolute risk for a meaningful time period | n/a |
| Other analyses | 17 | Report other analyses done—eg analyses of subgroups and interactions, and sensitivity analyses | Page 14 |
| **Discussion** | | |  |
| Key results | 18 | Summarise key results with reference to study objectives | Page 14 & 15 |
| Limitations | 19 | Discuss limitations of the study, taking into account sources of potential bias or imprecision. Discuss both direction and magnitude of any potential bias | Page 17 |
| Interpretation | 20 | Give a cautious overall interpretation of results considering objectives, limitations, multiplicity of analyses, results from similar studies, and other relevant evidence | Page 14 to 17 |
| Generalisability | 21 | Discuss the generalisability (external validity) of the study results | Page 15 & 16 |
| **Other information** | | |  |
| Funding | 22 | Give the source of funding and the role of the funders for the present study and, if applicable, for the original study on which the present article is based | Page 18 |

*Give information separately for exposed and unexposed groups.

**Note:** An Explanation and Elaboration article discusses each checklist item and gives methodological background and published examples of transparent reporting. The STROBE checklist is best used in conjunction with this article (freely available on the Web sites of PLoS Medicine at http://www.plosmedicine.org/, Annals of Internal Medicine at http://www.annals.org/, and Epidemiology at http://www.epidem.com/). Information on the STROBE Initiative is available at <http://www.strobe-statement.org>.

### Background

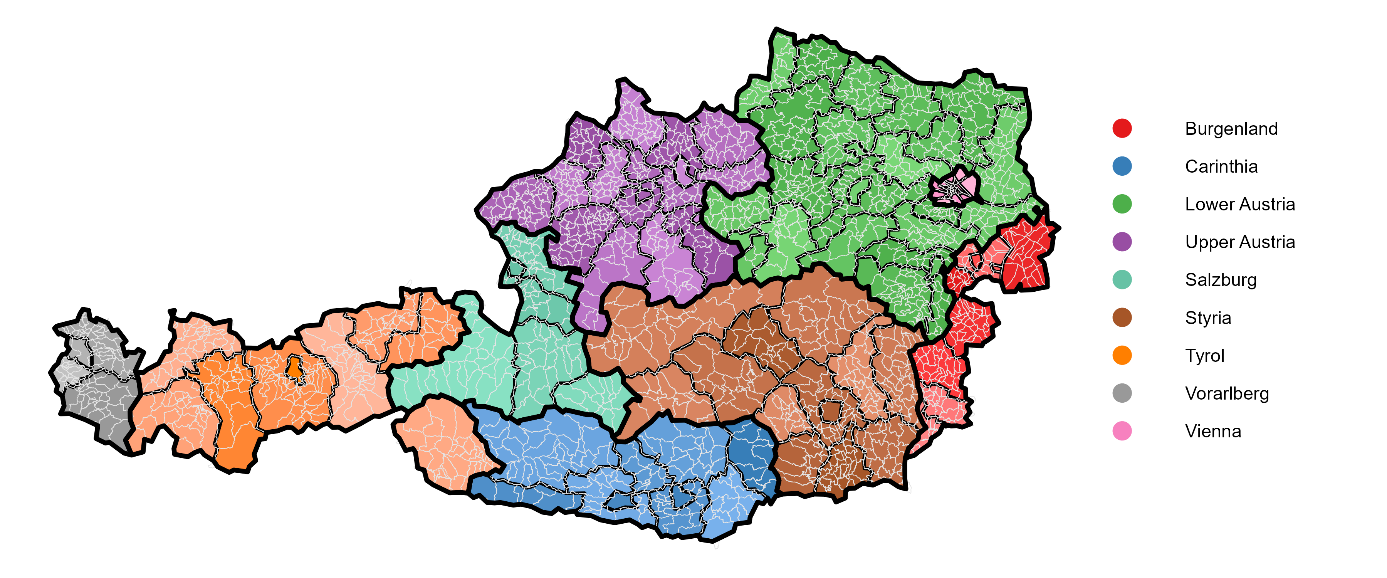

**Figure S1:** Map of Austria. States are colour coded and different districts separated by black lines (and slight variations on colour shading). Municipalities are indicated by white lines.

### Methods

##### Model convergence criteria and fallback procedure

A model fit was classified as converged only if it satisfied all of the following: (1) the Hessian of the marginal log-likelihood was positive definite, (2) the maximum absolute element of the gradient at convergence was below 0.01, and (3) standard errors were estimable (non-missing) for all fixed-effect coefficients in both the conditional (count) and zero-inflation components.

For the final primary (mortality) and secondary (infection) models, each year was fit independently and pooled across years by random-effects meta-analysis. When a year's model did not converge under the initial specification (negative binomial type 2 overdispersion [nbinom2] with the full zero-inflation formula), it was refit using a predetermined fallback sequence, applied in order until a converged fit was obtained:

1. **Warm start**: the same specification, with starting values for the fixed effects taken from the nearest previously converged year for that outcome and branch, rather than default starting values.
2. **Alternative overdispersion structure**: negative binomial type 1 (nbinom1), in which the variance increases linearly with the mean, in place of nbinom2.
3. **Zero-inflation formula simplification**: if the model still failed to converge, the zero-inflation component was simplified in a stepwise manner from the full specification (infection count, mean income, and log population) down to infection count and income, then infection count alone, then an intercept-only zero-inflation term, and finally no zero-inflation component (standard negative binomial). Each simplified zero-inflation specification was tried with both nbinom2 and nbinom1 before moving to the next simplification.

This fallback sequence was applied independently for each year and each variable-selection branch. A year that still failed to converge after exhausting all fallback options was excluded from the meta-analytic pooling for that specific outcome/branch/variable combination. The number of years contributing to each pooled estimate is reported alongside the results. The same convergence criteria and fallback logic were used in the descriptive univariable screening stage.

##### Municipality definition

As stated in the main manuscript, municipalities were defined as of January 1, 2022 [1]. This is relevant because on January 1, 2022 three municipalities merged. For datasets that were only available in the previous mapping, we summed the values of those three municipalities. This includes the pre-2022 entries in individual-level COVID-19 data. On January 1, 2025, two more municipalities combined into one (“Söchau” was merged with a now bigger “Fürstenfeld”). In cases where the relevant data was only available after January 1, 2025, we adapted Fürstenfeld to its old muniplicity number (62267) and manually added Söchau (62252). Urban/semi-rural/rural categorisation was only available for 2025 mapping. We manually defined Söchau as rural. Even though data for 2022 was available for percentage of 75+ year olds, the data was only available based on the 2025 municipality mapping. We used the same percentage for Söchau and Fürstenfeld that was stated for Fürstenfeld in the post-merger data mapping. For age category data we used ratio of total population to allocate the same ratio per age group to Söchau and Fürstenfeld.

##### Handling of Missing Values in UV data

Multiple days had missing values in the UV-satellite data. Visual inspection showed that days with more than 2% missing values were non-random (Figure S12), and interpolation attempts of those non-random missing data was deemed implausible. If less than 2% of the datapoints were missing on a given day missing values were interpolated using the R package “gapfill” [2]. To avoid biasing the averages due to missing values, UV-values for all days with 2% or more missing values were dropped from the analysis. Number of days that were used to estimate final municipality averages were in calendar years were: “2020”: 356 (missing: 10); “2021”: 342 (missing: 23); “2022”: 298 (missing: 66); and “2023”: 179 (missing: 2; note that “2023” ends on the 30^th^ of June). This is true for both UVA and UVB measures.

Calculation of Absolute Humidity

Absolute humidity (AH, g/m³) was derived from hourly gridded relative humidity (RH, %) and air temperature (T, °C) data. Saturation vapour pressure was calculated using the Magnus-Tetens approximation, following the parameterization of Sonntag (1990) [3]:

$$e_{s}=6.1078\times\exp\left( \frac{17.27T}{T+237.3} \right)$$

where *e*ₛ is saturation vapour pressure (hPa) and *T* is air temperature (°C).

Actual vapour pressure (*e*, hPa) was obtained by scaling saturation vapour pressure with relative humidity:

$$e=\left( \frac{RH}{100} \right)\times e_{s}$$

Absolute humidity (g/m³) was then calculated from the ideal gas law for water vapour:

$$AH=\frac{e\times2165}{T+273.15}$$

where *T* is expressed in °C, the term (*T* + 273.15) converts temperature to Kelvin, and 2165 is the specific gas constant term for water vapour expressed in units yielding AH in g/m³ (derived from the universal gas constant and the molar mass of water [4]). This calculation was applied independently to each hourly RH–temperature grid cell pair prior to any spatial or temporal aggregation

#### *Variables*

##### Temperature

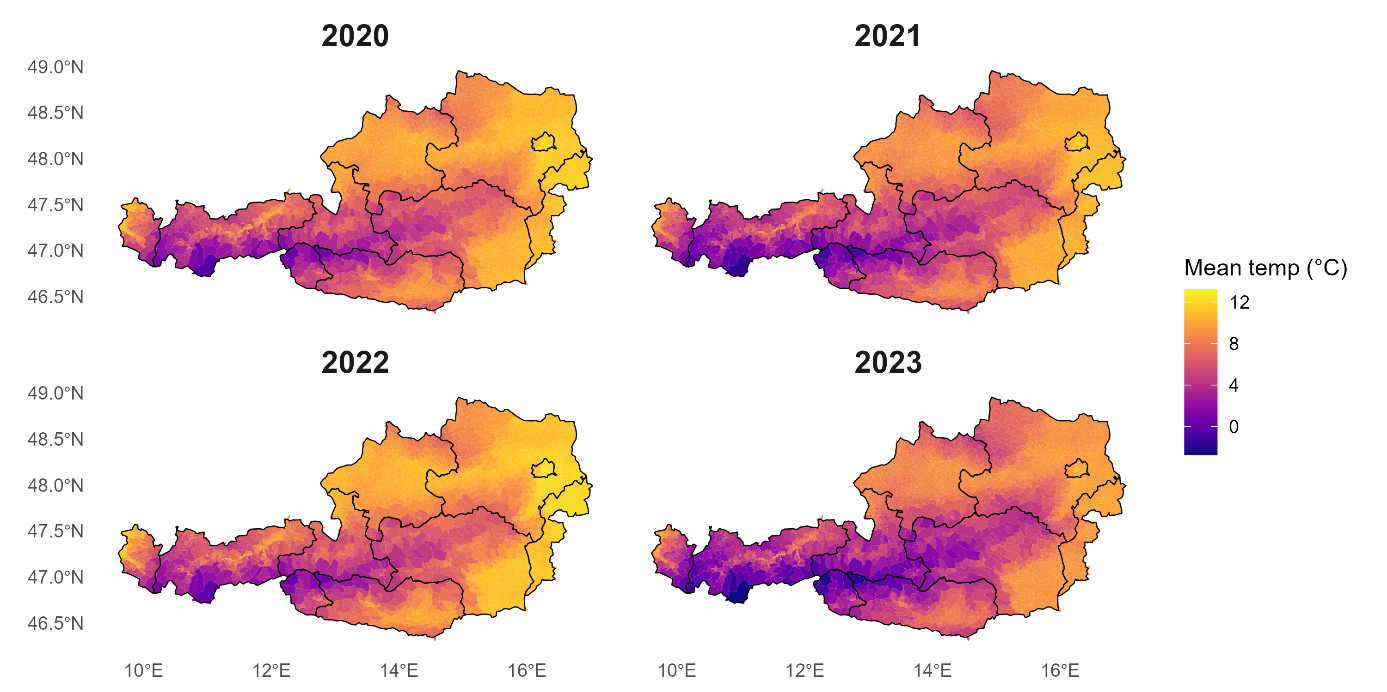

**Figure S2:** Average municipality temperature (°C) per observation period.

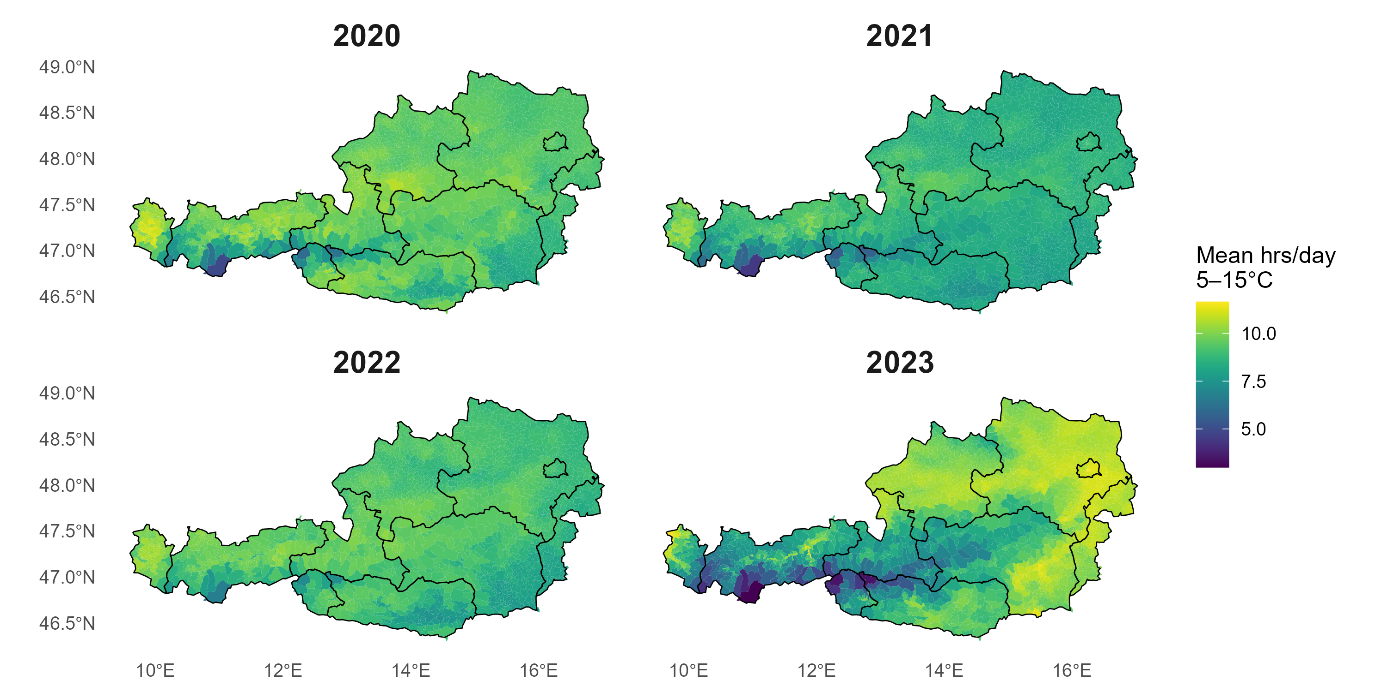

**Figure S3:** Mean hours per day with temperature between 5-15 °C per municipality and observation period.

##### Precipitation

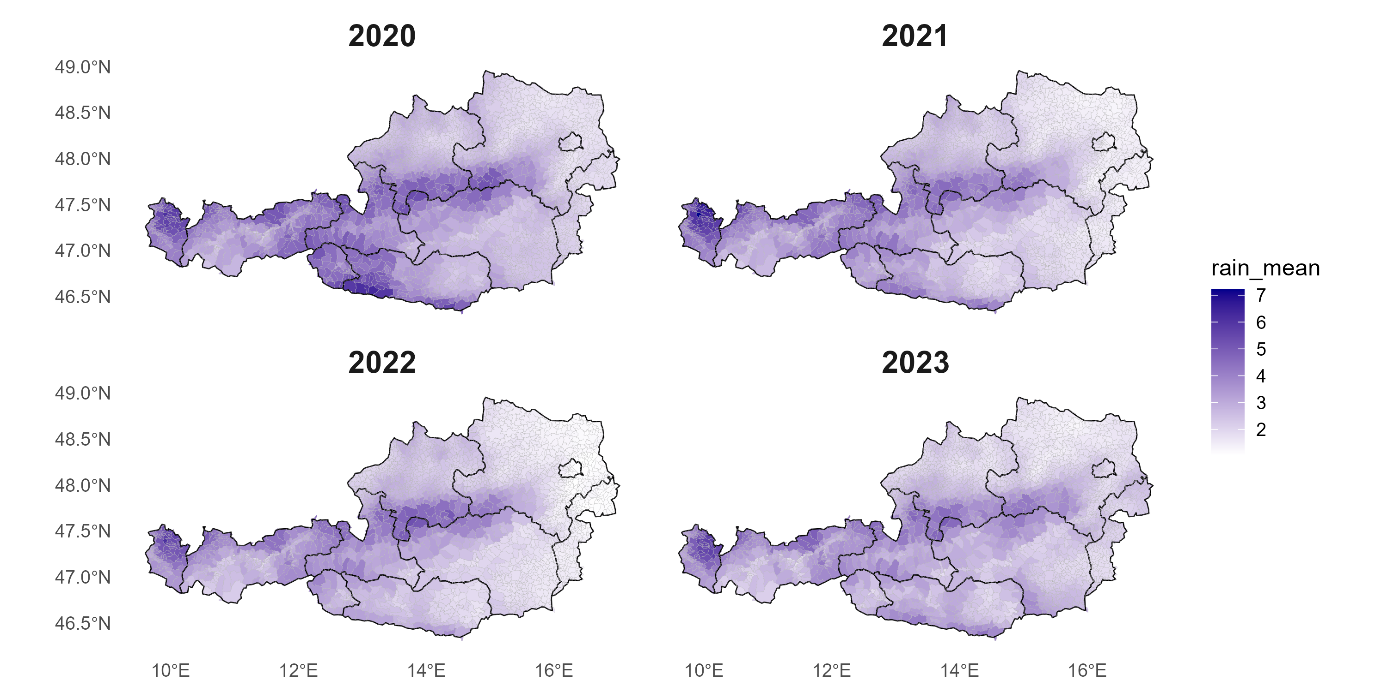

**Figure S4:** Average municipality precipitation (mm/day) per observation period.

##### Absolut Humidity

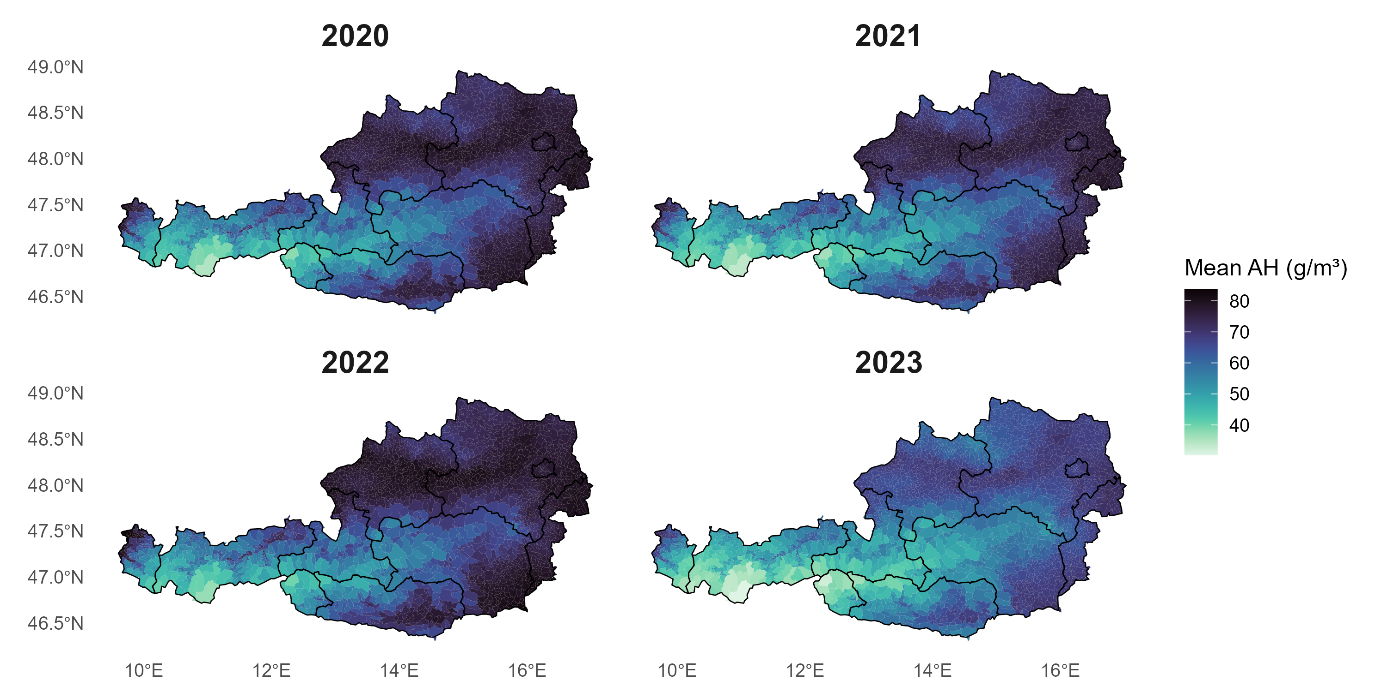
**Figure S5:** Average municipality absolute humidity (g/m³) per observation period.

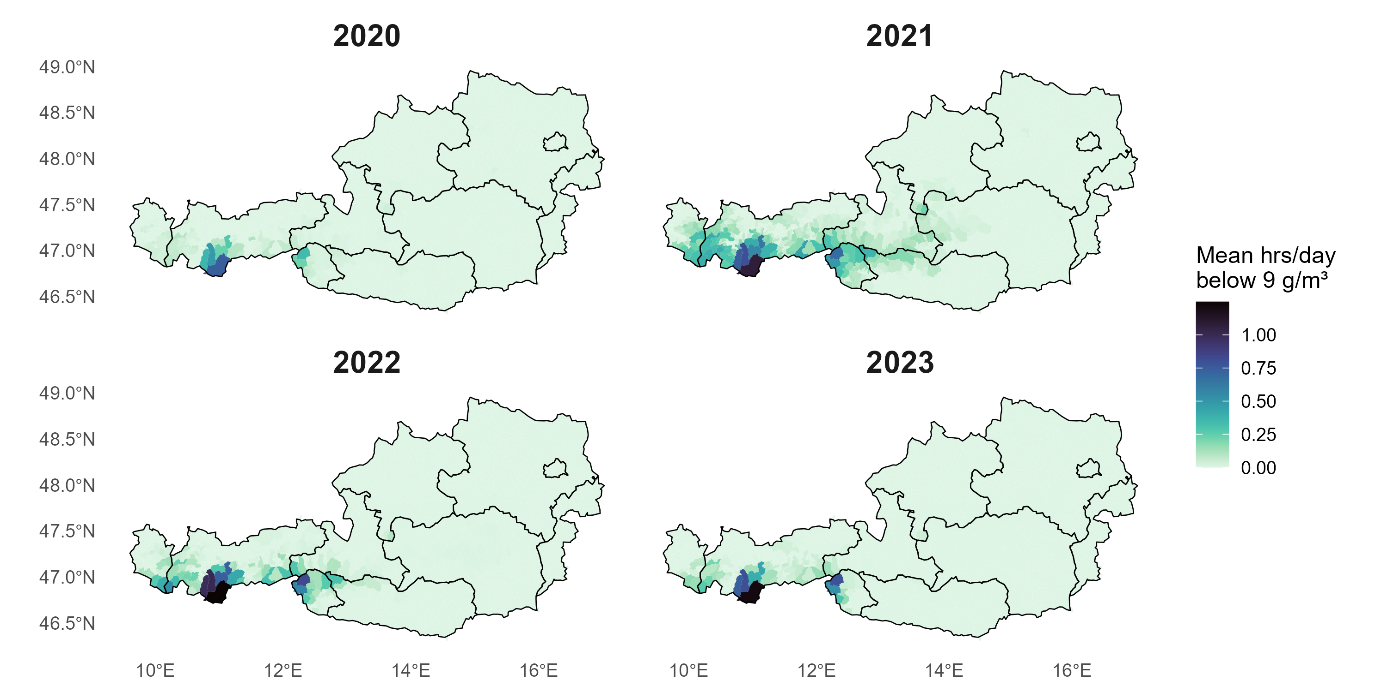
 **Figure S6:** Mean hours per day absolute humidity below 9 g/m³ per municipality and observation period.

##### Pollution

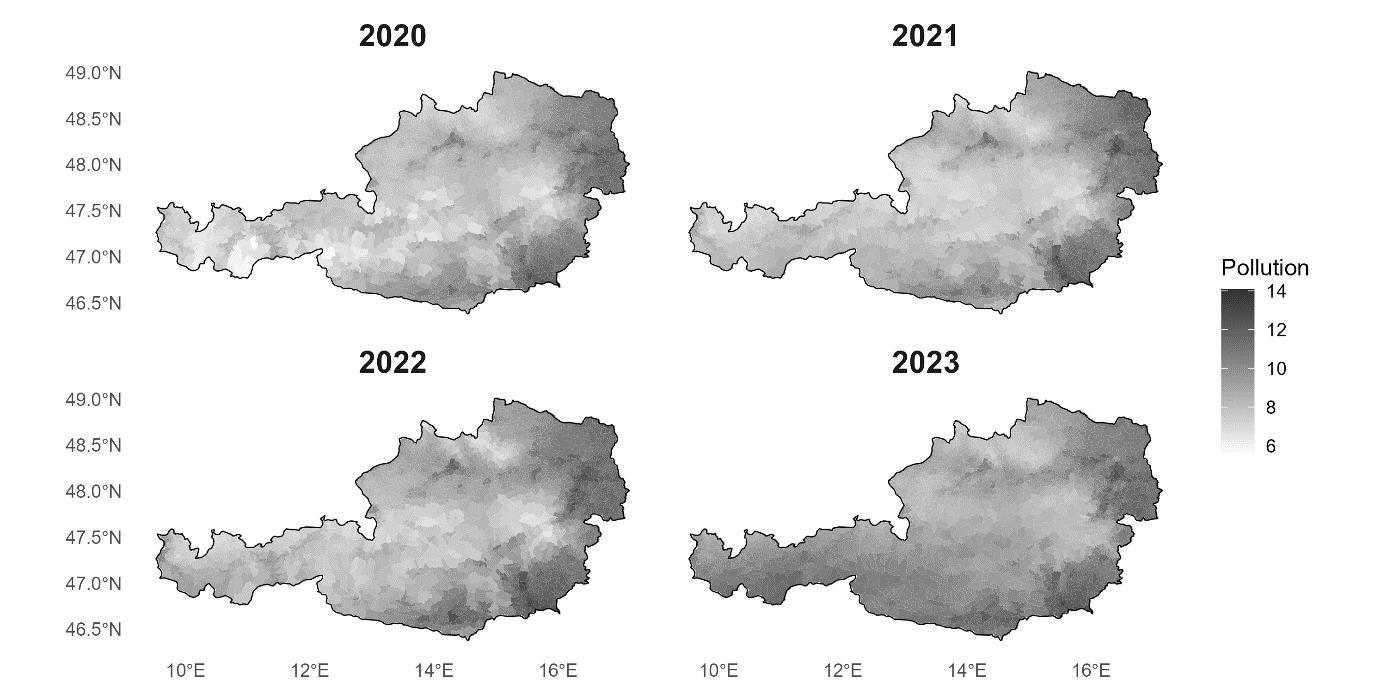
**Figure S7**: PM2.5 pollutions averages per town (µg/m³) for the respective observation periods (values from model version V6.GL.02.04 were used [5]).

##### Altitude

Data for altitude were available on 10m x 10m resolution. However, we analyzed an upscaled version of the altitude data (100m x 100m) to improve computational speed (Figure S8 and S9).

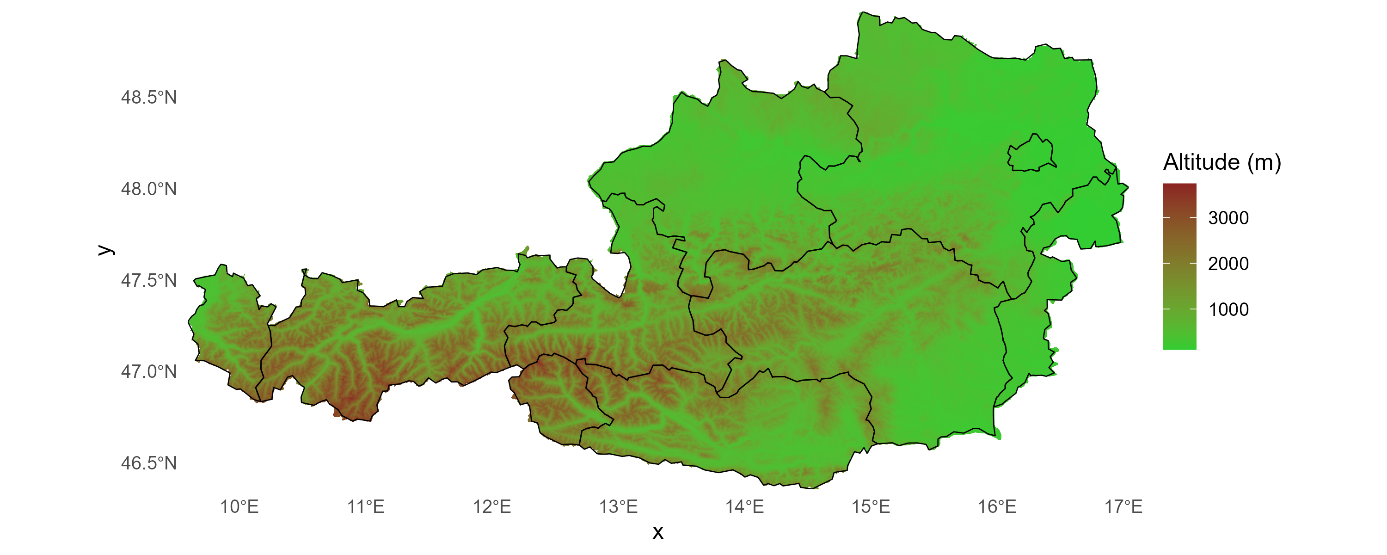

**Figure S8:** Raw raster data of altitude (m) in Austria up scaled to 100x100 meter (as used in the analysis).

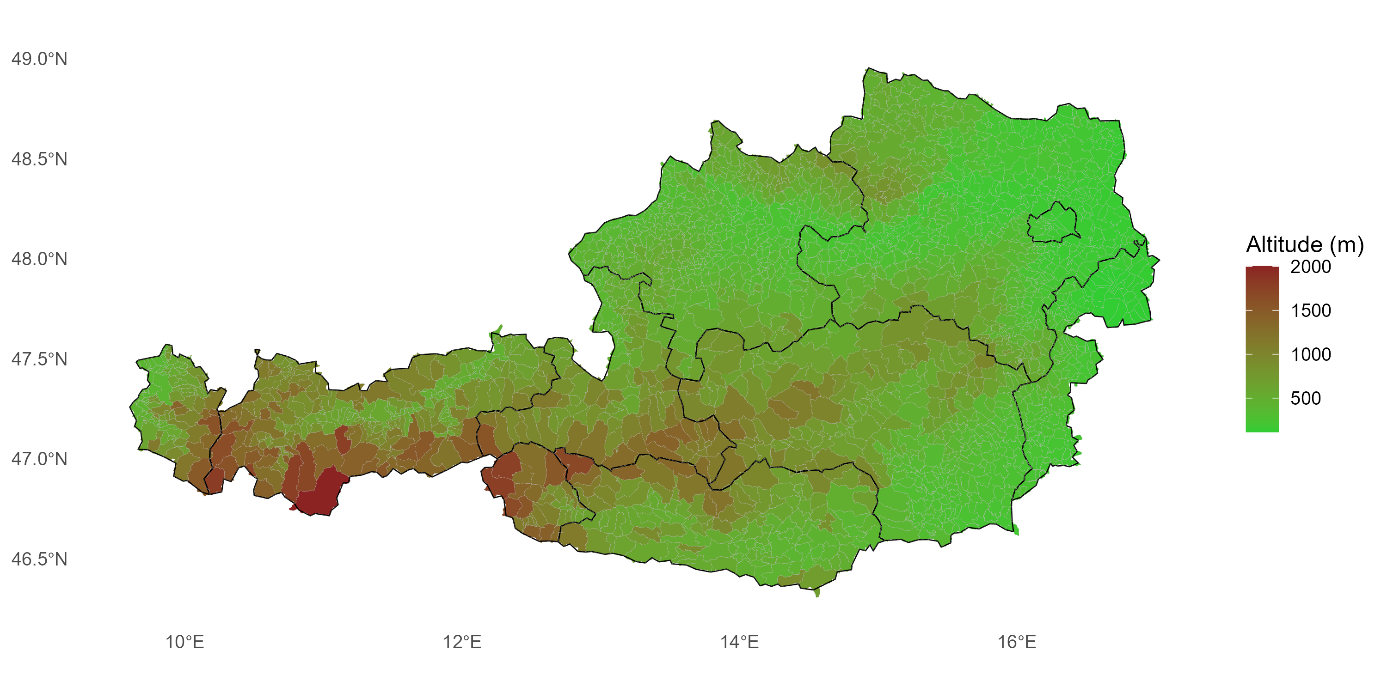
 **Figure S9:** The 10th percentile altitude (m)	value per municipality.

##### Ultraviolet radiation

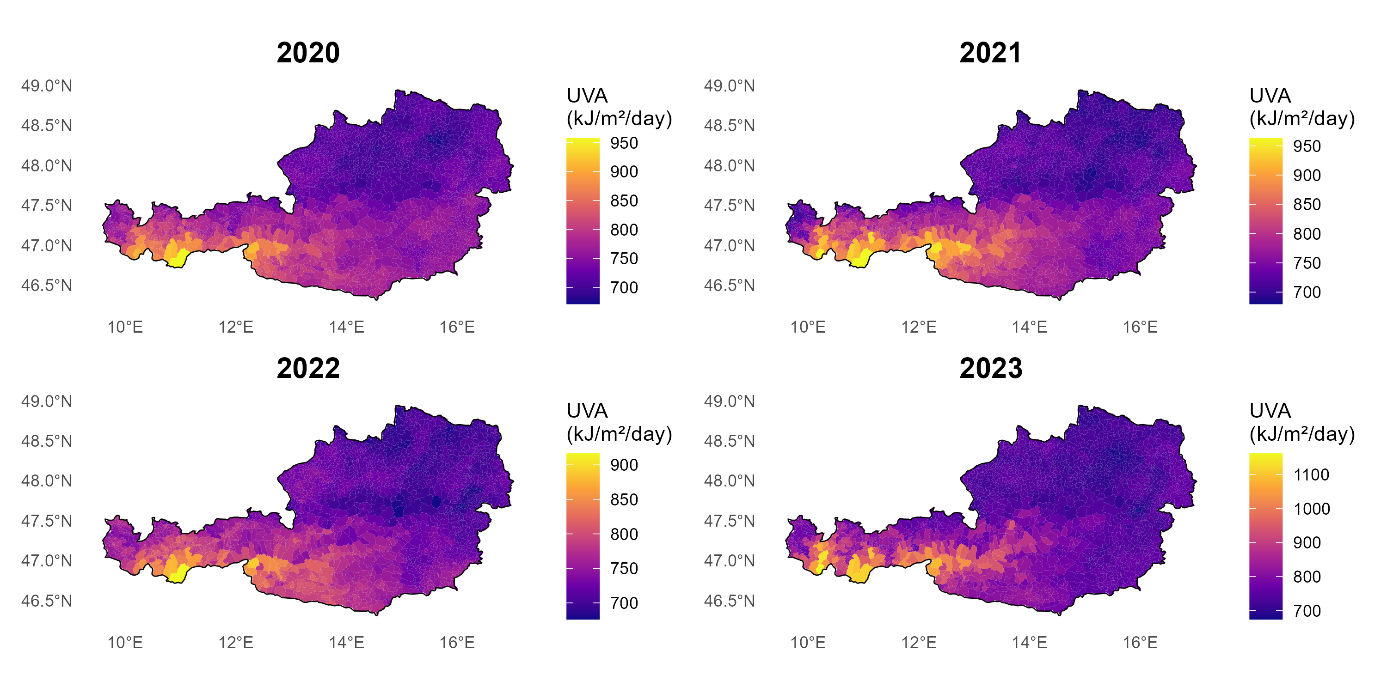
**Figure S10:** Average UVA value (KJ/m²/day) per municipality and observation period.

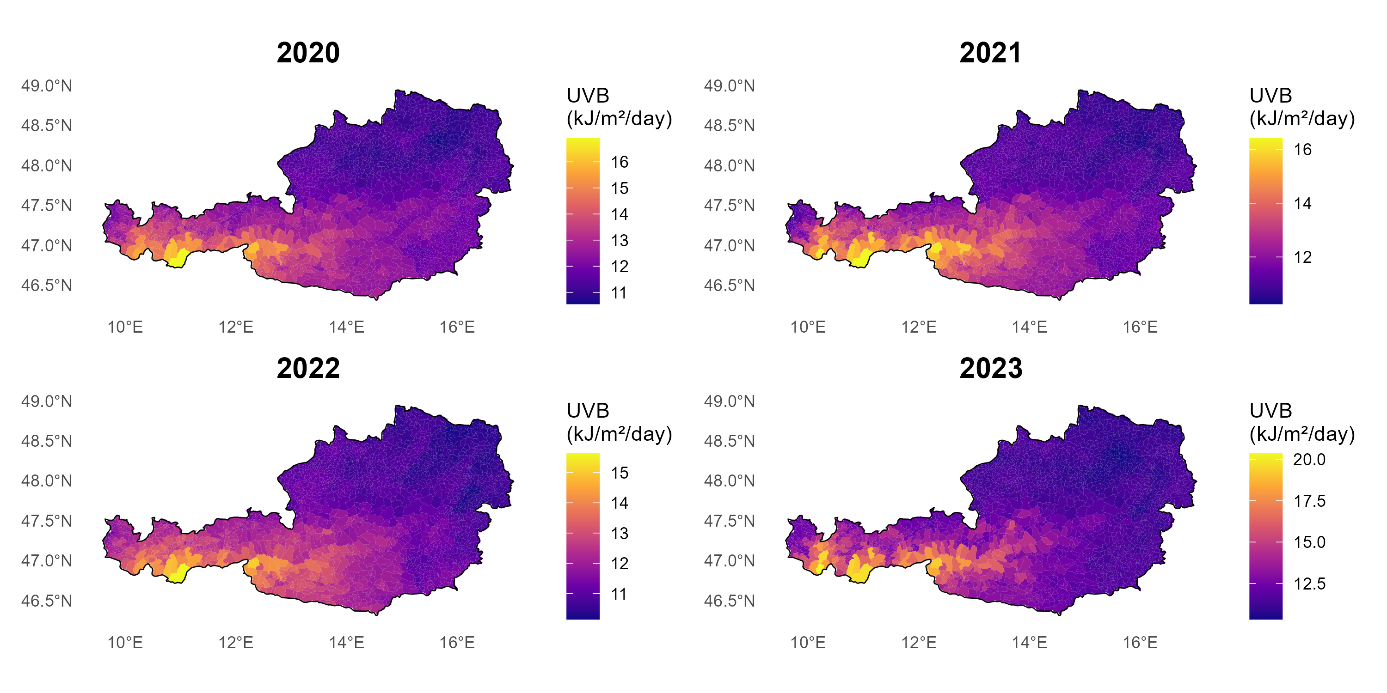

**Figure S11:** Average UVB value (KJ/m²/day) per municipality and observation period.

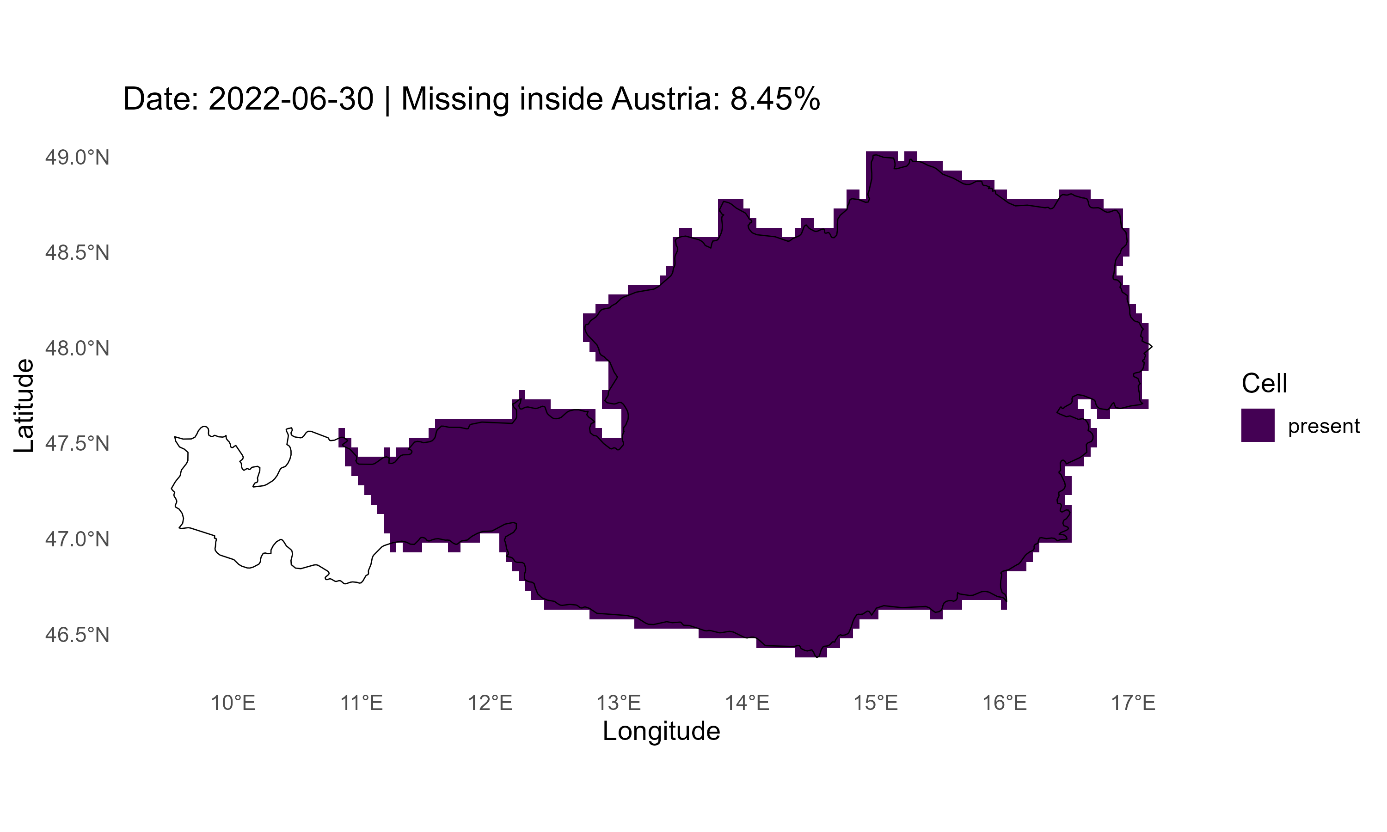
**Figure S12:** Example of daily UV estimates with more than 2% missing values. Note that missing values are not random and are thus generally not reliably estimated.

### Supplementary Results

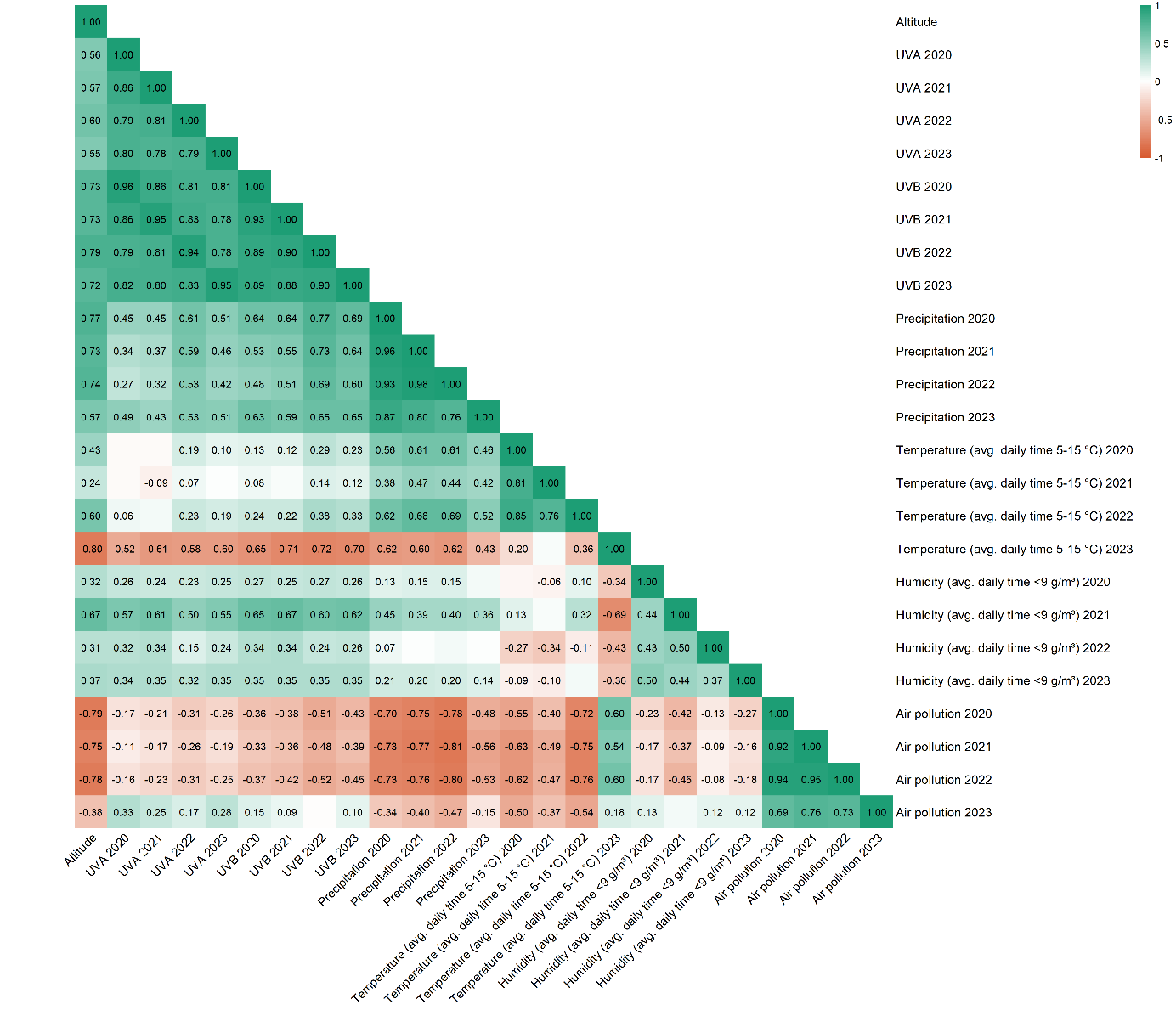

**Figure S13:** Heatmap of yearly environmental variables.

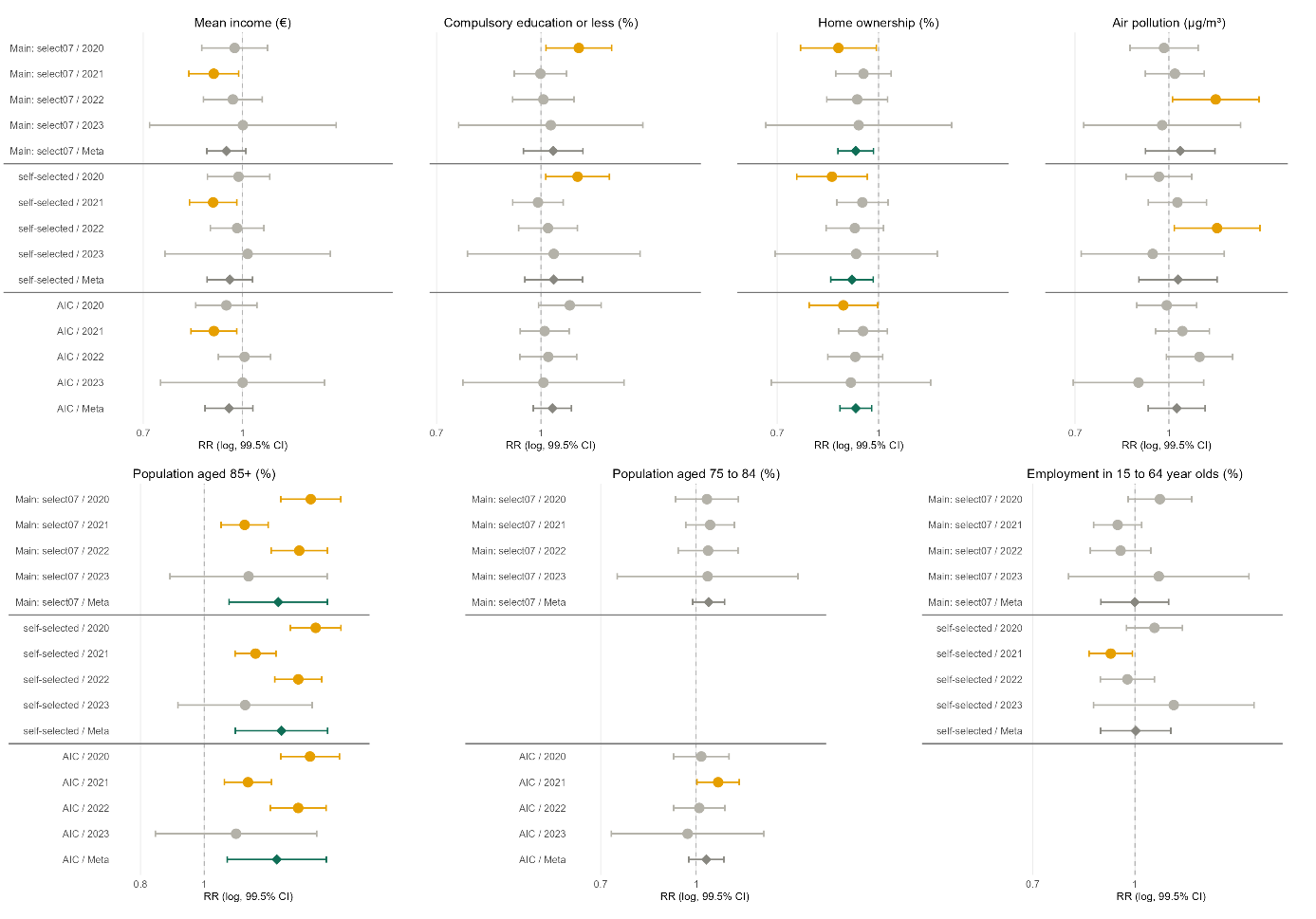

**Figure S14**: Comparison of different predictor choices: Multivariate yearly and meta-analysis outcomes for adjusted RRs of predictors of COVID-19 mortality with at least one significant estimate (p < 0.005).

| **Table S2: Univariable associations with COVID-19 deaths by year.** | | | | | | | | | |
| --- | --- | --- | --- | --- | --- | --- | --- | --- | --- |
| **Domain** | **Variable** | **2020** | | **2021** | | **2022** | | **2023** | |
|  |  | **RR (99.5% CI)** | **p** | **RR (99.5% CI)** | **p** | **RR (99.5% CI)** | **p** | **RR (99.5% CI)** | **p** |
| **Infections** | Infection count | 1.02 (0.96–1.08) | 0.34 | 1.00 (0.96–1.03) | 0.73 | 1.02 (0.99–1.05) | 0.11 | 1.06 (1.00–1.12) | 0.007 |
| **Infection dissemination** | Commuters (%) | **0.74 (0.68–0.82)** | **<0.001** | **0.80 (0.75–0.86)** | **<0.001** | **0.79 (0.73–0.85)** | **<0.001** | 0.95 (0.75–1.20) | 0.52 |
|  | Overnight stays p.c. | 0.98 (0.88–1.09) | 0.55 | 0.97 (0.87–1.08) | 0.40 | 0.89 (0.76–1.05) | 0.049 | 1.08 (0.86–1.36) | 0.32 |
|  | Population density (pop/km²) | 0.99 (0.93–1.04) | 0.43 | 0.98 (0.95–1.02) | 0.22 | 1.00 (0.97–1.04) | 0.74 | 1.04 (0.98–1.10) | 0.086 |
|  | Urbanisation category (rural/ semi-urban/ urban) | 1.29 (0.95–1.75) | 0.020 | 1.00 (0.80–1.26) | 0.97 | 1.25 (0.98–1.60) | 0.011 | 0.82 (0.38–1.77) | 0.48 |
|  | Work migration balance (ratio) | **1.16 (1.06–1.27)** | **<0.001** | 1.02 (0.97–1.06) | 0.24 | **1.05 (1.00–1.11)** | **0.004** | 1.01 (0.92–1.12) | 0.73 |
| **Demographic structure** | 65 to 74 year olds (%) | **1.20 (1.09–1.31)** | **<0.001** | **1.15 (1.07–1.24)** | **<0.001** | 1.08 (0.99–1.18) | 0.010 | 0.89 (0.70–1.12) | 0.14 |
|  | 75 to 84 year olds (%) | **1.32 (1.21–1.44)** | **<0.001** | **1.16 (1.09–1.24)** | **<0.001** | **1.22 (1.13–1.31)** | **<0.001** | 0.98 (0.80–1.20) | 0.82 |
|  | 85 or older (%) | **1.53 (1.41–1.65)** | **<0.001** | **1.24 (1.16–1.32)** | **<0.001** | **1.36 (1.26–1.46)** | **<0.001** | 1.03 (0.83–1.28) | 0.72 |
|  | Share of non-citizens (%) | 1.06 (0.98–1.15) | 0.037 | 0.99 (0.94–1.04) | 0.52 | 1.05 (0.98–1.11) | 0.037 | **1.19 (1.04–1.36)** | **<0.001** |
| **Socioeconomic status** | Business units p.c. | 1.00 (0.93–1.08) | 0.92 | 1.01 (0.96–1.06) | 0.72 | 0.99 (0.94–1.05) | 0.81 | 0.97 (0.85–1.11) | 0.53 |
|  | Compulsory education or less (%) | **1.29 (1.19–1.40)** | **<0.001** | **1.09 (1.02–1.16)** | **<0.001** | **1.11 (1.03–1.19)** | **<0.001** | 1.10 (0.92–1.32) | 0.12 |
|  | Employment in 15 to 64 year olds (%) | **0.87 (0.80–0.95)** | **<0.001** | **0.90 (0.85–0.96)** | **<0.001** | **0.86 (0.80–0.93)** | **<0.001** | 0.98 (0.79–1.21) | 0.74 |
|  | Home ownership (%) | **0.87 (0.81–0.94)** | **<0.001** | 0.97 (0.93–1.02) | 0.12 | **0.92 (0.87–0.97)** | **<0.001** | **0.85 (0.76–0.95)** | **<0.001** |
|  | Mean income (€) | **0.88 (0.81–0.96)** | **<0.001** | **0.91 (0.86–0.97)** | **<0.001** | 0.99 (0.92–1.06) | 0.68 | 0.97 (0.80–1.17) | 0.65 |
|  | Part-time employment rate (%) | 0.93 (0.85–1.02) | 0.028 | 0.96 (0.89–1.03) | 0.11 | 0.96 (0.88–1.05) | 0.23 | 1.02 (0.83–1.26) | 0.77 |
|  | University graduates (%) | 0.93 (0.87–1.01) | 0.010 | **0.94 (0.90–0.99)** | **0.001** | 1.02 (0.97–1.08) | 0.28 | 1.02 (0.90–1.16) | 0.66 |
| **Health status** | District level nursing care allowance recipients (%) | **1.21 (1.09–1.34)** | **<0.001** | **1.24 (1.14–1.35)** | **<0.001** | **1.15 (1.04–1.27)** | **<0.001** | 0.85 (0.65–1.10) | 0.069 |
| **Environment** | Air pollution (µg/m³) | 1.02 (0.92–1.13) | 0.54 | 1.01 (0.93–1.08) | 0.84 | **1.11 (1.02–1.21)** | **<0.001** | 1.04 (0.86–1.26) | 0.53 |
|  | Altitude (m) | 0.99 (0.88–1.11) | 0.75 | 1.01 (0.92–1.11) | 0.75 | 0.92 (0.83–1.03) | 0.040 | 0.88 (0.66–1.17) | 0.20 |
|  | Humidity (avg. daily time <9 g/m³) | 0.96 (0.86–1.07) | 0.30 | 1.00 (0.92–1.08) | 0.91 | 0.99 (0.90–1.09) | 0.72 | 1.03 (0.93–1.14) | 0.42 |
|  | Precipitation (mm/day) | 0.95 (0.85–1.06) | 0.21 | 0.97 (0.88–1.06) | 0.31 | **0.90 (0.82–1.00)** | **0.004** | 0.89 (0.68–1.16) | 0.22 |
|  | Temperature (avg. daily time 5-15 °C) | 0.95 (0.87–1.04) | 0.087 | 0.94 (0.87–1.01) | 0.023 | 0.94 (0.86–1.03) | 0.056 | 1.12 (0.87–1.45) | 0.21 |
|  | UVA radiation (KJ/m²/day) | 1.03 (0.92–1.16) | 0.41 | 1.00 (0.91–1.10) | 0.95 | **0.90 (0.81–0.99)** | **0.003** | 0.80 (0.59–1.07) | 0.031 |
|  | UVB radiation (KJ/m²/day) | 1.04 (0.92–1.16) | 0.40 | 1.01 (0.92–1.11) | 0.80 | **0.89 (0.80–0.99)** | **0.002** | 0.82 (0.61–1.10) | 0.056 |
| RR = rate ratio per 1 SD increase in the (standardised) predictor; for the categorical urbanisation variable rural is the reference level. 99.5% CI. | | | | | | | | | |
| Bold: p < 0.005. p.c. = per capita. | | | | | | | | | |

| **Table S3: Univariable associations of each predictor with COVID-19 deaths and SARS-CoV-2 infections (pooled across years).** | | | | | | | |
| --- | --- | --- | --- | --- | --- | --- | --- |
| **Domain** | **Variable** | **Deaths (primary analysis)** | | | **Infections (secondary analysis)** | | |
|  |  | **RR (99.5% CI)** | **p** | **I² (%)** | **RR (99.5% CI)** | **p** | **I² (%)** |
| **Infections** | Infection count | 1.02 (0.99–1.05) | 0.11 | 54 | – | – | – |
| **Infection dissemination** | Commuters (%) | **0.79 (0.73–0.86)** | **<0.001** | 62 | **1.04 (1.01–1.08)** | **0.001** | 95 |
|  | Overnight stays p.c. | 0.97 (0.91–1.03) | 0.16 | 0 | 0.99 (0.97–1.02) | 0.42 | 90 |
|  | Population density (pop/km²) | 1.00 (0.97–1.03) | 0.95 | 36 | 1.01 (0.95–1.08) | 0.54 | 98 |
|  | Urbanisation category (rural/ semi-urban/ urban) | 1.13 (0.91–1.42) | 0.11 | 50 | 1.01 (0.93–1.11) | 0.67 | 87 |
|  | Work migration balance (ratio) | 1.06 (0.97–1.15) | 0.055 | 84 | 1.01 (0.97–1.06) | 0.47 | 97 |
| **Demographic structure** | 65 to 74 year olds (%) | 1.09 (0.94–1.27) | 0.11 | 89 | 0.97 (0.93–1.00) | 0.007 | 94 |
|  | 75 to 84 year olds (%) | **1.18 (1.01–1.37)** | **0.002** | 91 | 0.98 (0.92–1.04) | 0.31 | 99 |
|  | 85 or older (%) | **1.29 (1.04–1.60)** | **0.001** | 96 | 1.00 (0.95–1.05) | 0.84 | 98 |
|  | Share of non-citizens (%) | 1.06 (0.96–1.17) | 0.096 | 85 | 0.99 (0.94–1.06) | 0.81 | 98 |
| **Socioeconomic status** | Business units p.c. | 1.00 (0.97–1.03) | 0.98 | 0 | 1.00 (0.99–1.01) | 0.64 | 34 |
|  | Compulsory education or less (%) | **1.15 (1.02–1.29)** | **0.001** | 87 | 1.00 (0.94–1.06) | 0.92 | 98 |
|  | Employment in 15 to 64 year olds (%) | **0.89 (0.85–0.93)** | **<0.001** | 5 | 1.02 (0.98–1.06) | 0.10 | 96 |
|  | Home ownership (%) | **0.91 (0.83–0.99)** | **0.001** | 85 | 0.99 (0.92–1.07) | 0.68 | 99 |
|  | Mean income (€) | 0.93 (0.86–1.01) | 0.017 | 70 | 1.01 (0.89–1.15) | 0.81 | 100 |
|  | Part-time employment rate (%) | 0.96 (0.91–1.00) | 0.007 | 0 | 1.01 (0.98–1.04) | 0.28 | 92 |
|  | University graduates (%) | 0.98 (0.91–1.05) | 0.31 | 74 | 1.01 (0.89–1.15) | 0.75 | 100 |
| **Health status** | District level nursing care allowance recipients (%) | 1.12 (0.90–1.38) | 0.15 | 93 | **0.95 (0.92–0.97)** | **<0.001** | 16 |
| **Environment** | Air pollution (µg/m³) | 1.04 (0.97–1.13) | 0.10 | 54 | 1.00 (0.90–1.12) | 0.95 | 99 |
|  | Altitude (m) | 0.97 (0.90–1.04) | 0.24 | 32 | 0.96 (0.83–1.12) | 0.48 | 99 |
|  | Humidity (avg. daily time <9 g/m³) | 0.99 (0.95–1.04) | 0.74 | 0 | 0.99 (0.97–1.02) | 0.60 | 93 |
|  | Precipitation (mm/day) | **0.94 (0.89–0.99)** | **0.002** | 0 | 0.98 (0.89–1.08) | 0.60 | 98 |
|  | Temperature (avg. daily time 5-15 °C) | **0.95 (0.90–0.99)** | **0.002** | 0 | 1.03 (0.93–1.14) | 0.37 | 99 |
|  | UVA radiation (KJ/m²/day) | 0.95 (0.83–1.08) | 0.28 | 77 | 0.98 (0.92–1.05) | 0.51 | 96 |
|  | UVB radiation (KJ/m²/day) | 0.95 (0.84–1.09) | 0.31 | 76 | 0.97 (0.88–1.08) | 0.46 | 98 |
| RR = rate ratio per 1 SD increase in the (standardised) predictor; for the categorical urbanisation variable rural is the reference level. 99.5% CI. | | | | | | | |
| Bold: p < 0.005. p.c. = per capita. | | | | | | | |
| I² = between-year heterogeneity of the pooled estimate. Years = number of years contributing to the pooled estimate. | | | | | | | |

| **Table S4: Multivariable associations with COVID-19 deaths by variable-selection approach.** | | | | | | | | | | |
| --- | --- | --- | --- | --- | --- | --- | --- | --- | --- | --- |
| **Domain** | **Variable** | **Main analysis (select07)** | | | **Sensitivity: self-selected** | | | **Sensitivity: select07 + AIC** | | |
|  |  | **RR (99.5% CI)** | **p** | **I² (%)** | **RR (99.5% CI)** | **p** | **I² (%)** | **RR (99.5% CI)** | **p** | **I² (%)** |
| **Infections** | Infection count | 1.00 (0.97–1.02) | 0.66 | 0 | 1.00 (0.98–1.02) | 0.98 | 0 | 1.00 (0.98–1.02) | 0.98 | 0 |
| **Infection dissemination** | Overnight stays p.c. | 1.01 (0.93–1.10) | 0.68 | 0 | – | – | – | – | – | – |
|  | Population density (pop/km²) | 1.00 (0.97–1.03) | 0.78 | 0 | 0.99 (0.97–1.02) | 0.42 | 0 | – | – | – |
|  | Urbanisation: Semi-urban (vs. reference) | 1.01 (0.86–1.17) | 0.90 | 9 | 1.01 (0.87–1.17) | 0.81 | 7 | – | – | – |
|  | Urbanisation: Urban (vs. reference) | 0.99 (0.89–1.11) | 0.88 | 0 | 1.01 (0.90–1.13) | 0.83 | 0 | – | – | – |
|  | Work migration balance (ratio) | 1.02 (0.97–1.07) | 0.23 | 11 | – | – | – | 1.02 (0.98–1.07) | 0.17 | 0 |
| **Demographic structure** | 65 to 74 year olds (%) | 0.97 (0.91–1.04) | 0.24 | 5 | – | – | – | – | – | – |
|  | 75 to 84 year olds (%) | 1.05 (0.99–1.11) | 0.027 | 0 | – | – | – | 1.04 (0.97–1.11) | 0.10 | 30 |
|  | 85 or older (%) | **1.30 (1.09–1.54)** | **<0.001** | 89 | **1.31 (1.12–1.54)** | **<0.001** | 91 | **1.29 (1.08–1.53)** | **<0.001** | 89 |
|  | Share of non-citizens (%) | 0.94 (0.87–1.01) | 0.012 | 0 | **0.93 (0.87–1.00)** | **0.003** | 0 | 0.94 (0.89–1.00) | 0.006 | 0 |
| **Socioeconomic status** | Business units p.c. | 0.97 (0.90–1.04) | 0.19 | 38 | – | – | – | 0.97 (0.91–1.03) | 0.16 | 31 |
|  | Compulsory education or less (%) | 1.04 (0.94–1.15) | 0.25 | 62 | 1.04 (0.95–1.15) | 0.22 | 62 | 1.04 (0.97–1.11) | 0.092 | 26 |
|  | Employment in 15 to 64 year olds (%) | 1.00 (0.89–1.12) | 0.99 | 72 | 1.00 (0.89–1.13) | 0.95 | 80 | – | – | – |
|  | Home ownership (%) | **0.92 (0.87–0.98)** | **<0.001** | 0 | **0.91 (0.85–0.98)** | **<0.001** | 32 | **0.92 (0.87–0.98)** | **<0.001** | 0 |
|  | Mean income (€) | 0.94 (0.88–1.01) | 0.020 | 24 | 0.96 (0.88–1.04) | 0.12 | 47 | 0.95 (0.87–1.04) | 0.11 | 54 |
|  | Part-time employment rate (%) | 1.02 (0.96–1.08) | 0.46 | 14 | – | – | – | – | – | – |
| **Health status** | District level nursing care allowance recipients (%) | 1.01 (0.92–1.11) | 0.75 | 28 | – | – | – | – | – | – |
| **Environment** | Air pollution (µg/m³) | 1.04 (0.92–1.19) | 0.36 | 64 | 1.04 (0.89–1.20) | 0.51 | 73 | 1.03 (0.92–1.15) | 0.44 | 61 |
|  | Humidity (avg. daily time <9 g/m³) | 1.01 (0.93–1.09) | 0.83 | 41 | 1.01 (0.93–1.10) | 0.79 | 61 | – | – | – |
|  | Precipitation (mm/day) | 0.98 (0.91–1.06) | 0.54 | 0 | 0.98 (0.91–1.06) | 0.46 | 0 | 0.96 (0.90–1.03) | 0.092 | 0 |
|  | Temperature (avg. daily time 5-15 °C) | 0.98 (0.91–1.06) | 0.46 | 25 | 0.98 (0.89–1.08) | 0.57 | 48 | – | – | – |
|  | UVA radiation (KJ/m²/day) | 0.95 (0.82–1.10) | 0.30 | 76 | 0.96 (0.84–1.08) | 0.30 | 66 | – | – | – |
| RR = rate ratio per 1 SD increase in the (standardised) predictor; for the categorical urbanisation variable rural is the reference level. 99.5% CI. | | | | | | | | | | |
| Bold: p < 0.005. p.c. = per capita. | | | | | | | | | | |
| I² = between-year heterogeneity of the pooled estimate. Years = number of years contributing to the pooled estimate. | | | | | | | | | | |

| **Table S5: Multivariable associations with COVID-19 deaths by year: main analysis.** | | | | | | | | | |
| --- | --- | --- | --- | --- | --- | --- | --- | --- | --- |
| **Domain** | **Variable** | **2020** | | **2021** | | **2022** | | **2023** | |
|  |  | **RR (99.5% CI)** | **p** | **RR (99.5% CI)** | **p** | **RR (99.5% CI)** | **p** | **RR (99.5% CI)** | **p** |
| **Infections** | Infection count | 0.99 (0.93–1.05) | 0.56 | 0.99 (0.96–1.03) | 0.70 | 0.99 (0.96–1.03) | 0.67 | 1.03 (0.95–1.13) | 0.28 |
| **Infection dissemination** | Overnight stays p.c. | 1.04 (0.91–1.19) | 0.39 | 0.97 (0.85–1.11) | 0.56 | 0.97 (0.80–1.17) | 0.61 | 1.22 (0.89–1.66) | 0.080 |
|  | Population density (pop/km²) | 0.99 (0.93–1.06) | 0.75 | 1.00 (0.95–1.05) | 0.96 | 0.99 (0.95–1.04) | 0.63 | 1.02 (0.91–1.15) | 0.61 |
|  | Urbanisation: Semi-urban (vs. reference) | 1.02 (0.76–1.36) | 0.88 | 0.94 (0.74–1.18) | 0.42 | 1.11 (0.87–1.41) | 0.22 | 0.78 (0.35–1.72) | 0.38 |
|  | Urbanisation: Urban (vs. reference) | 0.92 (0.73–1.16) | 0.31 | 1.06 (0.88–1.26) | 0.38 | 0.97 (0.79–1.20) | 0.72 | 1.01 (0.55–1.86) | 0.97 |
|  | Work migration balance (ratio) | 1.06 (0.96–1.16) | 0.10 | 0.99 (0.92–1.06) | 0.75 | 1.02 (0.95–1.11) | 0.39 | 1.09 (0.85–1.39) | 0.33 |
| **Demographic structure** | 65 to 74 year olds (%) | 1.00 (0.88–1.12) | 0.92 | 0.99 (0.90–1.09) | 0.87 | 0.92 (0.82–1.04) | 0.051 | 0.98 (0.69–1.39) | 0.87 |
|  | 75 to 84 year olds (%) | 1.04 (0.93–1.17) | 0.34 | 1.05 (0.96–1.15) | 0.11 | 1.05 (0.94–1.17) | 0.26 | 1.04 (0.74–1.46) | 0.72 |
|  | 85 or older (%) | **1.45 (1.31–1.61)** | **<0.001** | **1.15 (1.06–1.25)** | **<0.001** | **1.40 (1.26–1.54)** | **<0.001** | 1.17 (0.89–1.54) | 0.11 |
|  | Share of non-citizens (%) | 0.95 (0.82–1.09) | 0.26 | 0.93 (0.83–1.04) | 0.063 | 0.94 (0.83–1.07) | 0.18 | 0.99 (0.69–1.42) | 0.93 |
| **Socioeconomic status** | Business units p.c. | 0.94 (0.85–1.05) | 0.11 | 1.02 (0.94–1.10) | 0.57 | 0.95 (0.86–1.04) | 0.10 | 0.88 (0.65–1.20) | 0.25 |
|  | Compulsory education or less (%) | **1.14 (1.02–1.27)** | **0.001** | 1.00 (0.91–1.09) | 0.96 | 1.01 (0.91–1.12) | 0.83 | 1.03 (0.76–1.42) | 0.76 |
|  | Employment in 15 to 64 year olds (%) | 1.09 (0.98–1.22) | 0.027 | 0.94 (0.87–1.02) | 0.044 | 0.95 (0.86–1.06) | 0.18 | 1.09 (0.79–1.49) | 0.46 |
|  | Home ownership (%) | **0.87 (0.76–0.99)** | **0.003** | 0.95 (0.86–1.05) | 0.13 | 0.93 (0.83–1.03) | 0.049 | 0.93 (0.67–1.29) | 0.55 |
|  | Mean income (€) | 0.97 (0.86–1.09) | 0.50 | **0.90 (0.82–0.99)** | **0.001** | 0.97 (0.87–1.07) | 0.36 | 1.00 (0.72–1.40) | 0.99 |
|  | Part-time employment rate (%) | 1.07 (0.96–1.19) | 0.080 | 0.99 (0.91–1.08) | 0.68 | 1.01 (0.91–1.11) | 0.86 | 1.01 (0.76–1.35) | 0.89 |
| **Health status** | District level nursing care allowance recipients (%) | 1.02 (0.89–1.17) | 0.68 | 1.07 (0.95–1.21) | 0.100 | 0.97 (0.85–1.11) | 0.50 | 0.84 (0.59–1.20) | 0.17 |
| **Environment** | Air pollution (µg/m³) | 0.98 (0.86–1.12) | 0.69 | 1.02 (0.91–1.14) | 0.57 | **1.19 (1.01–1.41)** | **0.002** | 0.97 (0.72–1.31) | 0.81 |
|  | Humidity (avg. daily time <9 g/m³) | 0.92 (0.80–1.05) | 0.074 | 1.01 (0.92–1.11) | 0.79 | 1.04 (0.93–1.17) | 0.28 | 1.05 (0.91–1.21) | 0.34 |
|  | Precipitation (mm/day) | 0.95 (0.84–1.09) | 0.32 | 0.99 (0.87–1.12) | 0.83 | 1.01 (0.88–1.17) | 0.77 | 0.96 (0.69–1.34) | 0.75 |
|  | Temperature (avg. daily time 5-15 °C) | 0.94 (0.84–1.05) | 0.11 | 0.98 (0.88–1.09) | 0.60 | 1.04 (0.92–1.18) | 0.34 | 0.86 (0.54–1.36) | 0.35 |
|  | UVA radiation (KJ/m²/day) | 1.06 (0.94–1.20) | 0.19 | 0.96 (0.86–1.07) | 0.28 | 0.91 (0.82–1.02) | 0.019 | 0.66 (0.41–1.08) | 0.018 |
| RR = rate ratio per 1 SD increase in the (standardised) predictor; for the categorical urbanisation variable rural is the reference level. 99.5% CI. | | | | | | | | | |
| Bold: p < 0.005; p.c. = per capita. | | | | | | | | | |

| **Table S6: Variance explained by the multivariable models for each variable-selection approach.** | | | |
| --- | --- | --- | --- |
| **Outcome** | **Selection approach** | **R² marginal (retained predictors)** | **R² conditional (predictors + district random intercept)** |
| **Deaths (primary)** | Main analysis (select07) | 0.20 (0.07–0.29) | 0.31 (0.27–0.35) |
|  | Sensitivity: self-selected | 0.18 (0.06–0.28) | 0.31 (0.25–0.35) |
|  | Sensitivity: select07 + AIC | 0.17 (0.03–0.27) | 0.30 (0.23–0.36) |
| **Infections (secondary)** | Main analysis (select07) | 0.27 (0.13–0.43) | 0.63 (0.53–0.70) |
|  | Sensitivity: self-selected | 0.17 (0.05–0.23) | 0.61 (0.52–0.70) |
|  | Sensitivity: select07 + AIC | 0.20 (0.09–0.26) | 0.62 (0.51–0.69) |
| Values are proportions (0–1): mean across years, with the range across years in parentheses. | | | |
| Nakagawa R² on the log-rate scale (population offset excluded): marginal = proportion of variance explained by the retained predictors; conditional = predictors plus the district random intercept. | | | |

| **Table S7: Variance explained by year, outcome and variable-selection approach.** | | | | |
| --- | --- | --- | --- | --- |
| **Outcome** | **Selection approach** | **Year** | **R² marginal (retained predictors)** | **R² conditional (predictors + district random intercept)** |
| **Deaths (primary)** | Main analysis (select07) | 2020 | 0.29 | 0.35 |
|  |  | 2021 | 0.18 | 0.28 |
|  |  | 2022 | 0.25 | 0.35 |
|  |  | 2023 | 0.07 | 0.27 |
|  | Sensitivity: self-selected | 2020 | 0.28 | 0.35 |
|  |  | 2021 | 0.16 | 0.27 |
|  |  | 2022 | 0.24 | 0.35 |
|  |  | 2023 | 0.06 | 0.25 |
|  | Sensitivity: select07 + AIC | 2020 | 0.27 | 0.36 |
|  |  | 2021 | 0.15 | 0.26 |
|  |  | 2022 | 0.23 | 0.35 |
|  |  | 2023 | 0.03 | 0.23 |
| **Infections (secondary)** | Main analysis (select07) | 2020 | 0.13 | 0.53 |
|  |  | 2021 | 0.31 | 0.61 |
|  |  | 2022 | 0.43 | 0.70 |
|  |  | 2023 | 0.22 | 0.68 |
|  | Sensitivity: self-selected | 2020 | 0.05 | 0.52 |
|  |  | 2021 | 0.19 | 0.56 |
|  |  | 2022 | 0.23 | 0.70 |
|  |  | 2023 | 0.22 | 0.68 |
|  | Sensitivity: select07 + AIC | 2020 | 0.09 | 0.51 |
|  |  | 2021 | 0.26 | 0.58 |
|  |  | 2022 | 0.25 | 0.69 |
|  |  | 2023 | 0.21 | 0.68 |
| Values are proportions. | | | | |
| Nakagawa R² on the log-rate scale (population offset excluded): marginal = proportion of variance explained by the retained predictors; conditional = predictors plus the district random intercept. | | | | |

| **Table S8: Univariable associations with SARS-CoV-2 infections by year.** | | | | | | | | | |
| --- | --- | --- | --- | --- | --- | --- | --- | --- | --- |
| **Domain** | **Variable** | **2020** | | **2021** | | **2022** | | **2023** | |
|  |  | **RR (99.5% CI)** | **p** | **RR (99.5% CI)** | **p** | **RR (99.5% CI)** | **p** | **RR (99.5% CI)** | **p** |
| **Infection dissemination** | Commuters (%) | **1.03 (1.01–1.05)** | **<0.001** | **1.08 (1.06–1.10)** | **<0.001** | **1.05 (1.05–1.06)** | **<0.001** | 1.01 (0.99–1.04) | 0.097 |
|  | Overnight stays p.c. | 1.00 (0.98–1.02) | 0.87 | 0.99 (0.97–1.01) | 0.080 | **1.01 (1.00–1.02)** | **<0.001** | **0.97 (0.94–0.99)** | **<0.001** |
|  | Population density (pop/km²) | 1.00 (0.98–1.02) | 0.96 | **0.97 (0.95–0.99)** | **<0.001** | **1.01 (1.00–1.02)** | **0.004** | **1.08 (1.05–1.11)** | **<0.001** |
|  | Urbanisation category (rural/ semi-urban/ urban) | 1.03 (0.94–1.12) | 0.42 | 0.95 (0.88–1.02) | 0.040 | 0.99 (0.97–1.02) | 0.28 | **1.11 (1.01–1.22)** | **0.001** |
|  | Work migration balance (ratio) | 1.02 (1.00–1.04) | 0.006 | **0.97 (0.96–0.99)** | **<0.001** | 1.00 (1.00–1.01) | 0.038 | **1.05 (1.03–1.07)** | **<0.001** |
| **Demographic structure** | 65 to 74 year olds (%) | **0.96 (0.94–0.98)** | **<0.001** | **0.94 (0.92–0.96)** | **<0.001** | **0.97 (0.96–0.98)** | **<0.001** | 1.00 (0.97–1.03) | 0.95 |
|  | 75 to 84 year olds (%) | **0.98 (0.96–1.00)** | **0.003** | **0.93 (0.91–0.94)** | **<0.001** | **0.97 (0.97–0.98)** | **<0.001** | **1.04 (1.01–1.06)** | **<0.001** |
|  | 85 or older (%) | **1.02 (1.00–1.04)** | **0.001** | **0.95 (0.94–0.97)** | **<0.001** | **0.99 (0.98–0.99)** | **<0.001** | **1.03 (1.00–1.05)** | **<0.001** |
|  | Share of non-citizens (%) | 0.99 (0.97–1.01) | 0.32 | **0.95 (0.94–0.97)** | **<0.001** | **0.98 (0.97–0.99)** | **<0.001** | **1.06 (1.03–1.08)** | **<0.001** |
| **Socioeconomic status** | Business units p.c. | 1.00 (0.98–1.02) | 0.69 | 1.00 (0.98–1.02) | 0.83 | 1.01 (1.00–1.01) | 0.011 | 0.99 (0.97–1.01) | 0.23 |
|  | Compulsory education or less (%) | **1.05 (1.03–1.08)** | **<0.001** | 1.01 (0.99–1.03) | 0.059 | **0.97 (0.96–0.97)** | **<0.001** | **0.96 (0.94–0.98)** | **<0.001** |
|  | Employment in 15 to 64 year olds (%) | 1.02 (1.00–1.04) | 0.020 | **1.05 (1.03–1.07)** | **<0.001** | **1.03 (1.02–1.03)** | **<0.001** | 0.99 (0.96–1.01) | 0.097 |
|  | Home ownership (%) | 0.99 (0.97–1.01) | 0.059 | **1.05 (1.03–1.06)** | **<0.001** | **1.01 (1.00–1.01)** | **0.003** | **0.92 (0.90–0.94)** | **<0.001** |
|  | Mean income (€) | **0.96 (0.94–0.99)** | **<0.001** | **0.93 (0.91–0.94)** | **<0.001** | **1.03 (1.02–1.04)** | **<0.001** | **1.14 (1.11–1.16)** | **<0.001** |
|  | Part-time employment rate (%) | 0.98 (0.96–1.00) | 0.020 | 1.02 (1.00–1.03) | 0.024 | **1.02 (1.01–1.02)** | **<0.001** | **1.04 (1.01–1.06)** | **<0.001** |
|  | University graduates (%) | **0.97 (0.95–0.99)** | **<0.001** | **0.93 (0.91–0.94)** | **<0.001** | **1.02 (1.02–1.03)** | **<0.001** | **1.15 (1.12–1.18)** | **<0.001** |
| **Health status** | District level nursing care allowance recipients (%) | 0.93 (0.87–1.00) | 0.005 | 0.95 (0.89–1.01) | 0.016 | **0.96 (0.94–0.98)** | **<0.001** | **0.90 (0.80–1.00)** | **0.004** |
| **Environment** | Air pollution (µg/m³) | **0.96 (0.93–1.00)** | **0.003** | **0.92 (0.90–0.95)** | **<0.001** | **1.03 (1.02–1.04)** | **<0.001** | **1.11 (1.07–1.15)** | **<0.001** |
|  | Altitude (m) | 1.02 (0.99–1.06) | 0.079 | **1.05 (1.02–1.08)** | **<0.001** | **0.97 (0.96–0.98)** | **<0.001** | **0.83 (0.79–0.86)** | **<0.001** |
|  | Humidity (avg. daily time <9 g/m³) | 1.00 (0.99–1.02) | 0.53 | 1.01 (0.99–1.03) | 0.11 | 1.00 (0.99–1.01) | 0.98 | **0.96 (0.94–0.99)** | **<0.001** |
|  | Precipitation (mm/day) | 1.01 (0.97–1.04) | 0.69 | **1.06 (1.03–1.10)** | **<0.001** | **0.97 (0.95–0.98)** | **<0.001** | **0.90 (0.87–0.93)** | **<0.001** |
|  | Temperature (avg. daily time 5-15 °C) | 1.00 (0.97–1.02) | 0.83 | 1.01 (0.99–1.03) | 0.14 | **0.98 (0.97–0.99)** | **<0.001** | **1.15 (1.11–1.19)** | **<0.001** |
|  | UVA radiation (KJ/m²/day) | 1.01 (0.97–1.05) | 0.53 | 1.02 (0.99–1.05) | 0.031 | 0.99 (0.98–1.01) | 0.10 | **0.92 (0.88–0.95)** | **<0.001** |
|  | UVB radiation (KJ/m²/day) | 1.02 (0.98–1.07) | 0.12 | 1.03 (1.00–1.06) | 0.008 | **0.97 (0.95–0.98)** | **<0.001** | **0.88 (0.85–0.91)** | **<0.001** |
| RR = rate ratio per 1 SD increase in the (standardised) predictor; for the categorical urbanisation variable rural is the reference level. 99.5% CI. | | | | | | | | | |
| Bold: p < 0.005; p.c. = per capita. | | | | | | | | | |

| **Table S9: Multivariable associations with SARS-CoV-2 infections by variable-selection approach.** | | | | | | | | | | |
| --- | --- | --- | --- | --- | --- | --- | --- | --- | --- | --- |
| **Domain** | **Variable** | **Main analysis (select07)** | | | **Sensitivity: self-selected** | | | **Sensitivity: select07 + AIC** | | |
|  |  | **RR (99.5% CI)** | **p** | **I² (%)** | **RR (99.5% CI)** | **p** | **I² (%)** | **RR (99.5% CI)** | **p** | **I² (%)** |
| **Infection dissemination** | Overnight stays p.c. | 1.00 (0.98–1.02) | 0.89 | 84 | – | – | – | 1.00 (0.98–1.02) | 0.85 | 81 |
|  | Population density (pop/km²) | 1.00 (0.99–1.01) | 0.96 | 0 | 1.01 (1.00–1.01) | 0.009 | 0 | 1.01 (1.00–1.01) | 0.021 | 0 |
|  | Urbanisation: Semi-urban (vs. reference) | 0.99 (0.97–1.01) | 0.11 | 0 | 0.99 (0.97–1.02) | 0.53 | 16 | – | – | – |
|  | Urbanisation: Urban (vs. reference) | 1.02 (0.97–1.07) | 0.33 | 78 | 1.02 (0.97–1.07) | 0.30 | 79 | – | – | – |
|  | Work migration balance (ratio) | 1.01 (1.00–1.02) | 0.061 | 60 | – | – | – | 1.01 (0.99–1.03) | 0.082 | 78 |
| **Demographic structure** | 65 to 74 year olds (%) | 0.98 (0.94–1.02) | 0.11 | 94 | – | – | – | 0.98 (0.94–1.02) | 0.12 | 95 |
|  | 75 to 84 year olds (%) | 0.98 (0.95–1.01) | 0.027 | 88 | – | – | – | 0.98 (0.95–1.02) | 0.26 | 96 |
|  | 85 or older (%) | 1.01 (0.97–1.05) | 0.40 | 96 | 1.00 (0.95–1.05) | 0.98 | 98 | – | – | – |
|  | Share of non-citizens (%) | 0.99 (0.96–1.01) | 0.14 | 81 | **0.99 (0.98–1.00)** | **<0.001** | 5 | 0.98 (0.96–1.00) | 0.017 | 68 |
| **Socioeconomic status** | Business units p.c. | 1.00 (0.98–1.02) | 0.96 | 74 | – | – | – | – | – | – |
|  | Compulsory education or less (%) | 1.01 (0.96–1.06) | 0.60 | 96 | 1.00 (0.95–1.06) | 0.81 | 97 | 1.01 (0.95–1.07) | 0.63 | 97 |
|  | Employment in 15 to 64 year olds (%) | **1.01 (1.01–1.02)** | **<0.001** | 0 | **1.02 (1.01–1.03)** | **<0.001** | 24 | **1.01 (1.00–1.02)** | **<0.001** | 0 |
|  | Home ownership (%) | 0.99 (0.94–1.04) | 0.55 | 95 | 0.98 (0.94–1.03) | 0.31 | 94 | 0.98 (0.93–1.04) | 0.45 | 96 |
|  | Mean income (€) | 1.01 (0.92–1.11) | 0.68 | 99 | 1.01 (0.92–1.11) | 0.76 | 99 | 1.02 (0.93–1.11) | 0.61 | 99 |
|  | Part-time employment rate (%) | **1.01 (1.00–1.02)** | **<0.001** | 2 | – | – | – | – | – | – |
| **Health status** | District level nursing care allowance recipients (%) | **0.97 (0.95–0.98)** | **<0.001** | 0 | – | – | – | – | – | – |
| **Environment** | Air pollution (µg/m³) | 0.99 (0.93–1.05) | 0.62 | 94 | 0.99 (0.93–1.05) | 0.57 | 94 | 0.99 (0.93–1.04) | 0.45 | 93 |
|  | Humidity (avg. daily time <9 g/m³) | 1.00 (0.99–1.01) | 0.69 | 8 | 1.00 (0.99–1.01) | 0.75 | 11 | 1.00 (0.99–1.01) | 0.55 | 58 |
|  | Precipitation (mm/day) | 0.99 (0.95–1.02) | 0.26 | 82 | 0.98 (0.95–1.02) | 0.14 | 76 | 0.99 (0.95–1.03) | 0.45 | 86 |
|  | Temperature (avg. daily time 5-15 °C) | 1.00 (0.98–1.02) | 0.75 | 59 | 1.00 (0.98–1.03) | 0.56 | 66 | 1.01 (0.98–1.04) | 0.30 | 84 |
|  | UVA radiation (KJ/m²/day) | 0.99 (0.97–1.01) | 0.24 | 55 | 0.99 (0.98–1.00) | 0.047 | 3 | – | – | – |
| RR = rate ratio per 1 SD increase in the (standardised) predictor; for the categorical urbanisation variable rural is the reference level. 99.5% CI. | | | | | | | | | | |
| Bold: p < 0.005. p.c. = per capita. | | | | | | | | | | |
| I² = between-year heterogeneity of the pooled estimate. Years = number of years contributing to the pooled estimate. | | | | | | | | | | |

| **Table S10: Multivariable associations with SARS-CoV-2 infections by year: main analysis.** | | | | | | | | | |
| --- | --- | --- | --- | --- | --- | --- | --- | --- | --- |
| **Domain** | **Variable** | **2020** | | **2021** | | **2022** | | **2023** | |
|  |  | **RR (99.5% CI)** | **p** | **RR (99.5% CI)** | **p** | **RR (99.5% CI)** | **p** | **RR (99.5% CI)** | **p** |
| **Infection dissemination** | Overnight stays p.c. | 1.01 (0.98–1.03) | 0.50 | **0.98 (0.96–1.00)** | **0.002** | **1.01 (1.01–1.02)** | **<0.001** | 1.00 (0.97–1.04) | 0.74 |
|  | Population density (pop/km²) | 1.00 (0.98–1.03) | 0.69 | 0.99 (0.97–1.01) | 0.31 | 1.00 (0.99–1.01) | 0.96 | 1.01 (0.98–1.04) | 0.59 |
|  | Urbanisation: Semi-urban (vs. reference) | 1.03 (0.94–1.12) | 0.44 | 1.01 (0.94–1.08) | 0.75 | 0.98 (0.96–1.01) | 0.053 | 0.98 (0.90–1.08) | 0.59 |
|  | Urbanisation: Urban (vs. reference) | 1.05 (0.98–1.12) | 0.047 | 1.05 (1.00–1.10) | 0.006 | 0.99 (0.97–1.00) | 0.011 | 1.00 (0.94–1.06) | 0.94 |
|  | Work migration balance (ratio) | **1.03 (1.00–1.06)** | **0.003** | 1.01 (0.99–1.03) | 0.36 | **1.01 (1.00–1.02)** | **<0.001** | 1.00 (0.97–1.02) | 0.61 |
| **Demographic structure** | 65 to 74 year olds (%) | **0.96 (0.94–0.99)** | **<0.001** | **0.95 (0.93–0.97)** | **<0.001** | **0.98 (0.98–0.99)** | **<0.001** | 1.02 (0.99–1.04) | 0.11 |
|  | 75 to 84 year olds (%) | **0.97 (0.95–1.00)** | **0.004** | **0.96 (0.94–0.98)** | **<0.001** | **0.98 (0.97–0.99)** | **<0.001** | 1.01 (0.98–1.04) | 0.42 |
|  | 85 or older (%) | **1.03 (1.01–1.06)** | **<0.001** | **0.97 (0.96–0.99)** | **<0.001** | **1.01 (1.00–1.01)** | **<0.001** | **1.03 (1.01–1.06)** | **<0.001** |
|  | Share of non-citizens (%) | **0.96 (0.93–1.00)** | **0.002** | 0.98 (0.96–1.01) | 0.073 | **0.98 (0.97–0.99)** | **<0.001** | 1.02 (0.98–1.05) | 0.19 |
| **Socioeconomic status** | Business units p.c. | 0.99 (0.96–1.01) | 0.20 | 1.02 (1.00–1.04) | 0.009 | 1.00 (0.99–1.00) | 0.21 | 0.99 (0.97–1.02) | 0.45 |
|  | Compulsory education or less (%) | **1.06 (1.03–1.09)** | **<0.001** | 1.02 (1.00–1.05) | 0.007 | **0.99 (0.98–1.00)** | **<0.001** | 0.97 (0.94–1.00) | 0.011 |
|  | Employment in 15 to 64 year olds (%) | 1.01 (0.99–1.04) | 0.18 | 1.01 (0.99–1.03) | 0.078 | **1.01 (1.00–1.02)** | **<0.001** | 1.02 (0.99–1.05) | 0.036 |
|  | Home ownership (%) | 1.00 (0.97–1.04) | 0.99 | 1.03 (1.00–1.05) | 0.008 | 0.99 (0.99–1.00) | 0.049 | **0.94 (0.90–0.97)** | **<0.001** |
|  | Mean income (€) | 1.00 (0.97–1.03) | 0.90 | **0.94 (0.92–0.96)** | **<0.001** | **1.02 (1.02–1.03)** | **<0.001** | **1.10 (1.07–1.14)** | **<0.001** |
|  | Part-time employment rate (%) | 1.00 (0.97–1.02) | 0.62 | 1.01 (0.99–1.03) | 0.074 | **1.01 (1.00–1.02)** | **<0.001** | 1.02 (0.99–1.05) | 0.022 |
| **Health status** | District level nursing care allowance recipients (%) | 0.94 (0.88–1.01) | 0.018 | 0.97 (0.92–1.03) | 0.15 | **0.97 (0.95–0.99)** | **<0.001** | 0.96 (0.89–1.05) | 0.23 |
| **Environment** | Air pollution (µg/m³) | **0.96 (0.91–1.00)** | **0.004** | **0.95 (0.92–0.98)** | **<0.001** | 1.01 (1.00–1.02) | 0.030 | **1.04 (1.00–1.08)** | **0.003** |
|  | Humidity (avg. daily time <9 g/m³) | 0.99 (0.97–1.02) | 0.47 | 1.01 (0.99–1.03) | 0.20 | 1.00 (0.99–1.01) | 0.72 | 0.99 (0.97–1.01) | 0.19 |
|  | Precipitation (mm/day) | 1.00 (0.95–1.04) | 0.77 | 1.01 (0.97–1.05) | 0.42 | 0.99 (0.97–1.00) | 0.006 | **0.95 (0.91–0.99)** | **<0.001** |
|  | Temperature (avg. daily time 5-15 °C) | 1.00 (0.97–1.03) | 0.72 | 1.01 (0.98–1.04) | 0.25 | 0.99 (0.98–1.00) | 0.006 | 1.02 (0.97–1.08) | 0.20 |
|  | UVA radiation (KJ/m²/day) | 1.01 (0.96–1.06) | 0.63 | 1.01 (0.97–1.04) | 0.58 | **0.98 (0.97–1.00)** | **<0.001** | 0.96 (0.92–1.01) | 0.030 |
| RR = rate ratio per 1 SD increase in the (standardised) predictor; for the categorical urbanisation variable rural is the reference level. 99.5% CI. | | | | | | | | | |
| Bold: p < 0.005; p.c. = per capita. | | | | | | | | | |
